# Calcium Channel Blockers Increased the Risk of Aortic Aneurysm and Dissection

**DOI:** 10.1101/2025.05.19.25327784

**Authors:** Tianfeng Ma, Zeyu Cai, Xinming Xu, Long Cao, Ao Wang, Zhongshuang Zhang, Siting Zhang, Zhenkun Huang, Jingjing Luo, Sen Lin, Xinhao Wang, Yi Fu, Fang Yu, Jing Zhou, Lixin Wang, Hongpeng Zhang, Xiang Gao, Wei Guo, Wei Kong

## Abstract

Aortic aneurysm and dissection (AAD) are life-threatening conditions for which there is a lack of effective pharmacological therapies. Hypertension is a known risk factor for AAD, leading to the common prescription of antihypertensive medications for AAD patients. The impaired contractility of vascular smooth muscle cells (VSMCs) is strongly linked to AAD, yet the role of calcium channel blockers (CCBs), which directly inhibit VSMC contractility, in AAD onset of hypertensive patients remains unclear. We thus analyzed data from 501,878 initially AAD-free participants and reported that CCB use had greater risks of AAD (adjusted HR=1.31, 95% CI: 1.17-1.48), TAAD (adjusted HR=1.23, 95% CI: 1.00-1.50), and AAA (adjusted HR=1.32, 95% CI: 1.13-1.53), than hypertensive patients not receiving antihypertensive medication. during a median follow-up of 13.5 years (*P*<0.001). In mouse models induced by angiotensin II, elastase, and β-aminopropionitrile (BAPN), various subtypes of CCBs significantly increased aortic stiffness and the risk of AAD. Of 95 patients with type B aortic dissection included after endovascular repair surgery in the secondary data analysis, CCBs (69 patients) limited aortic aneurysm/dissection regression compared with that associated with other antihypertensives (26 patients). Moreover, the silencing of protein kinase cGMP-dependent 1 (PRKG1) to restore VSMCs contractility significantly mitigated CCB-induced aortic stiffness and the incidence of AAD. These findings suggest that CCBs may increase AAD risk and post-stent surgery prognosis, highlighting the need for caution when prescribing CCBs to hypertensive patients at risk for AAD.

## Introduction

Aortic aneurysm and dissection (AAD) are life-threatening cardiovascular conditions associated with high mortality rates. Aortic aneurysm refers to the irreversible enlargement of the aorta, resulting in a diameter exceeding 1.5 times that of adjacent normal vessels. Aortic dissection is defined as disruption of the intima leading to separation of the aortic wall layers followed by the formation of a false lumen^1^. Depending on the location, AAD are classified into thoracic aortic aneurysm/dissection (TAAD) and abdominal aortic aneurysm (AAA)^2,3^. Although recognized as distinct disease entities, TAAD and AAA share common pathological changes, including vascular extracellular matrix degradation, inflammation, and dysfunction of vascular smooth muscle cells (VSMCs). Currently, the management of AAD relies solely on surgical intervention, including artificial blood vessel replacement and endovascular stent graft repair^4^. However, there are no currently proven drugs to slow aortic expansion or prevent aortic rupture in AAD patients (except for Marfan Syndrome), including blood pressure-lowering drugs^5–7^, lipid-modifying medications^8^, anti-inflammatory or antibiotics drugs^9–11^, anti-thrombotic drugs^12^.

Hypertension has long been recognized as an independent risk factor for AAD^13,14^. In clinical practice, antihypertensive drugs, including angiotensin II receptor blockers (ARBs), calcium channel blockers (CCBs), and β-blockers, are commonly prescribed to AAD patients to reduce pressure on the aortic wall, but the effect of various antihypertensive drugs on AAD patients remains unclear. Notably, emerging evidence indicates that the development of AAD is attributed to the impaired VSMC contractility^15^. Reduced expression of contractile apparatus proteins (e.g., myosin heavy chain [MYH11], smooth muscle α-actin [SM α-actin] and smooth muscle protein 22-α [SM22-α]) is observed in the aortas of patients with TAAD and AAA^16^. Additionally, genetic mutations in myosin heavy chain 11 (*MYH11*), actin α 2 (*ACTA2*), myosin light chain kinase (*MYLK*), and protein kinase, cGMP-dependent, type I (*PRKG1*) are linked to TAAD^17,18^. Moreover, patients with AAD have shown reduced vascular contractility, accompanied by changes in mechanical properties, such as increased stiffness^19–21^. This raises concerns about whether antihypertensive drugs that inhibit VSMC contractility, such as CCBs^22^, pose potential risks to AAD onset. Previous studies have reported that CCBs (amlodipine, diltiazem, and azelnidipine), at doses 5-10 times higher than the clinical doses, may alleviate angiotensin II (AngII)-induced AAD formation when co-administered with AngII in mice^23–26^. However, a single study revealed that amlodipine and verapamil promoted aorta expansion in mice with Marfan syndrome (MFS)^27^. Furthermore, in a randomized controlled trial involving 224 patients with small AAAs, amlodipine failed to delay aortic expansion^7^. Currently, however, all clinical studies have focused on the medication treatment of patients with diagnosed AAD, while the impact of CCB use in hypertensive patients on the incidence of AAD remains unclear.

In this study, our objective was to evaluate the impact of CCBs on the risk of AAD. Here we report results of (1) the association between CCBs and their subtypes with subsequent AAD occurrence, using data from the UK Biobank; and (2) the effects of CCBs on both murine models and post-stent AAD patients.

## Results

### Association of CCB or Subtype Use with the Risk of AAD in UKB Participants

We first analyzed the association between CCBs and the incidence of AAD using data from the UK Biobank (Figure 1a). Among the 501,878 participants initially free of AAD, 28.3% were identified as having hypertension (Extended Data Table 1). These hypertensive participants were divided into three groups based on their medication use: 8.7% were not taking any antihypertensives, and 6.8% were using CCBs, either exclusively (5.7%) or in combination with other antihypertensive medications (1.1%). The remaining 12.8% were classified as using antihypertensive medications other than CCBs (Extended Data Tables 2-3). The baseline characteristics of the participants are shown in Extended Data Table 4. Hypertensive participants, regardless of medication use, were generally older, had higher body mass index (BMI), and higher systolic blood pressure (SBP) than those without hypertension. Among hypertensive groups, CCB users were older, had more men, fewer white participants, and with lower total cholesterol levels than untreated individuals or those using other antihypertensive medications.

**Figure 1.**
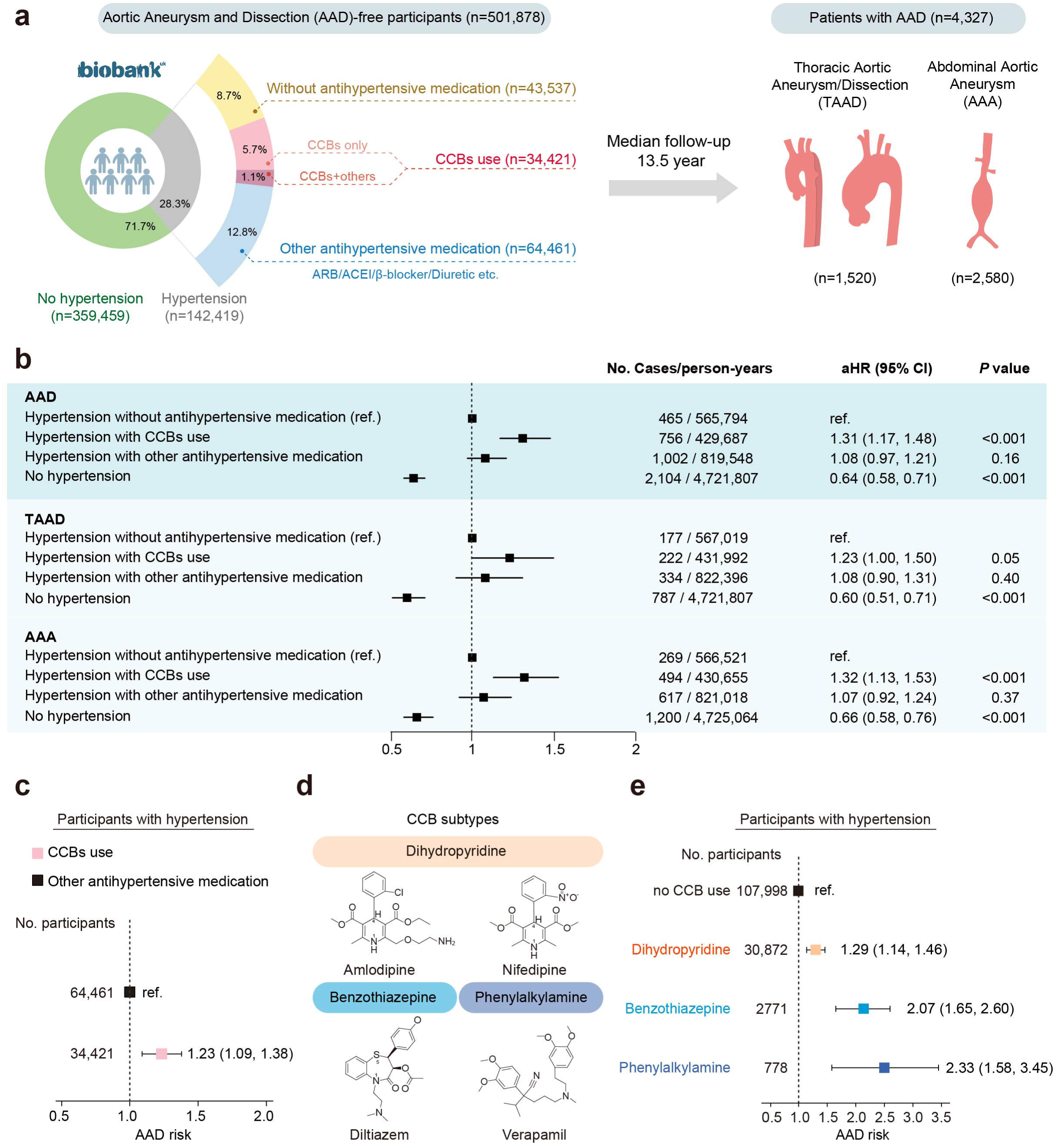
CCBs use and AAD/AAD subtypes risks in the UKB. **a**, Overview of the included AAD-free participants from the UKB (n=501,878). Pie chart showed the exposure groups categorized by hypertension and antihypertensive medications in relation to the total number of included participants. **b**, Association of hypertension status and antihypertensive medication use (CCBs or other antihypertensive medication excluding CCBs use) with AAD/AAD subtypes risks among the AAD-free UKB participants (n=501,878). **c**, Association of CCBs with AAD risk among participants with hypertension and any antihypertensive medication use (n=98,882, including 34,421 participants with hypertension using CCBs and 64,461 participants with hypertension using other antihypertensive medication use except for CCBs). **d**, Subtypes and chemical structures of CCBs **e**, Association of CCBs subtypes use with AAD risk among the UKB participants with hypertension (n=142,419). b, c, e, Adjusted hazard ratios of AAD risk were calculated via Cox model adjusted for age (years), sex (men; women), BMI (kg/m2), total cholesterol (mmol/L), triglycerides (mmol/L), smoking status (never; previous; current), education (College or University degree; A levels or equivalent; O levels or equivalent; None of the above), ethnicity (white; or others), systolic blood pressure (mmHg), and simultaneous use of other antihypertensive medications. AAD, Aortic aneurysm and dissection; TAAD, Thoracic aortic aneurysm and dissection; AAA, Abdominal aortic aneurysm; CCB, Calcium channel blocker; BMI, body mass index.

During a median follow-up period of 13.5 years, 4,327 cases of AAD were identified, including 1,520 TAAD and 2,580 AAA (Figure 1a and Extended Data Table 5). As expected, participants without hypertension had significantly lower risks of AAD, TAAD, and AAA compared to those with hypertension but without antihypertensive medication (*P*<0.001 for all) (Figure 1b). We further found that participants with hypertension and concurrent CCB use had greater risks of AAD (adjusted HR=1.31, 95% CI: 1.17-1.48), TAAD (adjusted HR=1.23, 95% CI: 1.00-1.50), and AAA (adjusted HR=1.32, 95% CI: 1.13-1.53), than hypertensive patients not receiving antihypertensive medication (Figure 1b). To minimize the indication bias that medication use itself might reflect disease severity, we restricted the analyses to the participants receiving antihypertensive medications and observed that CCB users still had a significantly greater risk of AAD (HR=1.23, 95% CI: 1.09-1.38), than other antihypertensive medication users (Figure 1c and Extended Data Table 6). Furthermore, among the participants with prevalent AAD, CCB use was associated with increased mortality (HR=1.52, 95% CI: 1.04-2.21) (Extended Data Table 7).

CCBs are generally divided into three main subtypes based on their chemical structure and action mechanisms: dihydropyridines, benzothiazepines and phenylalkylamines^28^(Figure 1d). All subtypes of CCBs were associated with a significantly greater AAD risk than no CCB use among participants with hypertension, with phenylalkylamine CCBs presenting the highest risk (HR=2.33, 95% CI: 1.58-3.45), benzothiazepine CCBs showing an intermediate risk (HR=2.07, 95% CI: 1.65-2.60), and dihydropyridine CCBs having a relatively lower risk (HR=1.29, 95% CI: 1.14-1.46), after further adjusting for concurrent use of other blood pressure medications (Figure 1e). The sensitivity analysis considering the competing risk of death, with the broader definition of hypertension, excluding coronary artery diseases, adjusting for eGFR or removing participants with hypertension combined with both concurrent use of CCBs and other antihypertensive medications, and yielded the similar results, supporting the robustness of our study (Extended Data Tables 8-12).

### CCBs Aggravated Both Thoracic and Abdominal Aortic Aneurysm Formation in AngII-infused mouse models

We next investigated the cause-effect of different CCB subtypes on the development of aortic aneurysms in an angiotensin II (AngII)-infused mouse model (Figure 2a). Sixteen-week-old C57BL/6J mice were subcutaneously infused with AngII (1000 ng/kg·min^-1^) via minipumps for a total of 28 days. Unlike previous studies^23–26^, which administered CCBs either simultaneously with or prior to AngII infusion, this study initiated CCB treatment 7 days after AngII infusion, once hypertension had been successfully induced. Mice that developed aortic aneurysms by Day 7 of AngII infusion, as detected by ultrasound, were excluded from further experiments. The mice were then treated daily for 21 days with either a vehicle or one of four CCBs: amlodipine (1 mg/kg/day), nifedipine (20 mg/kg/day), diltiazem (8 mg/kg/day) or verapamil (12 mg/kg/day). Saline-infused mice were treated with vehicle and served as a negative control. As expected, all four CCB treatments significantly reduced SBP in the AngII-infused mice (Figure 2b).

**Figure 2.**
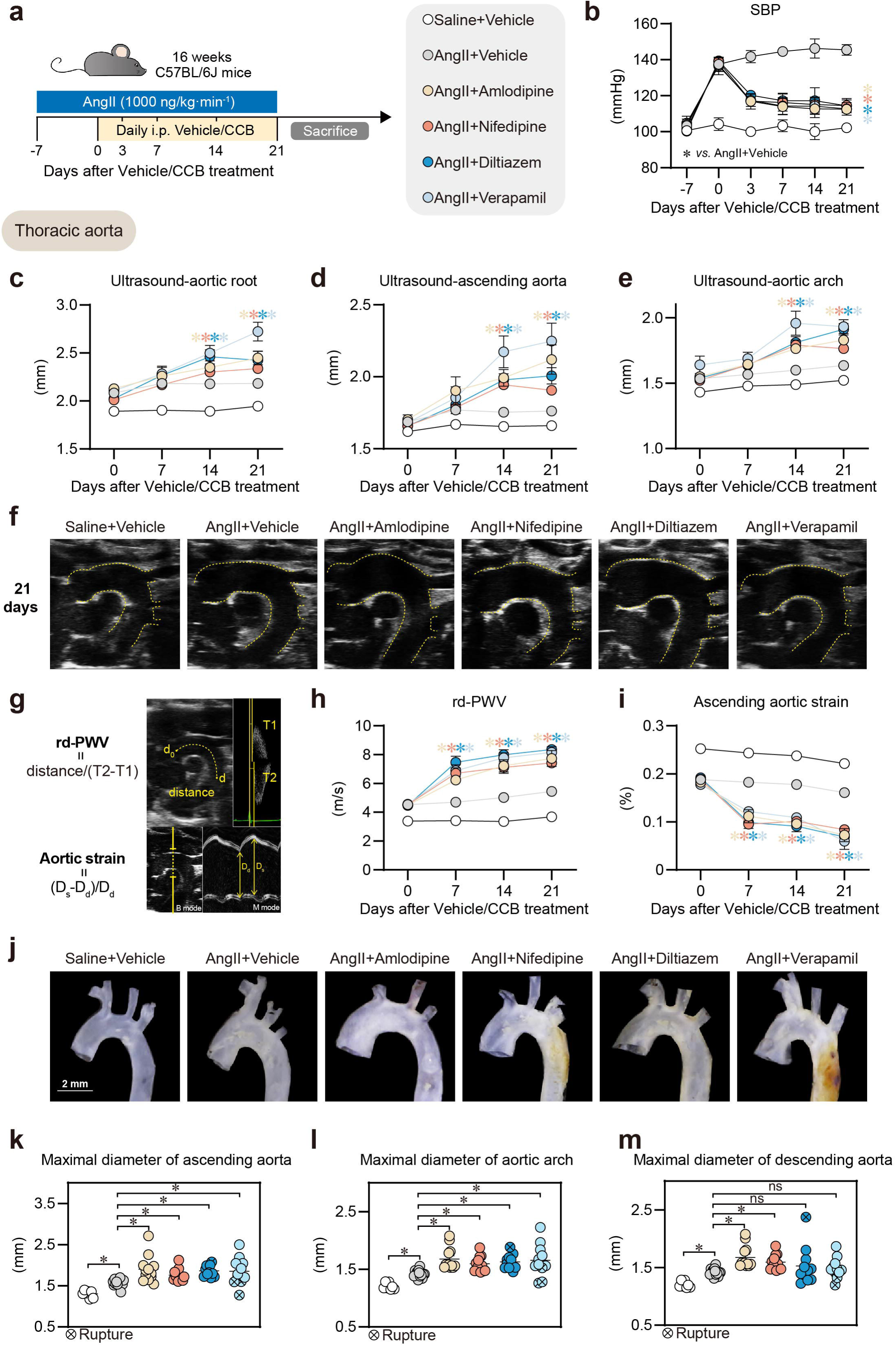
CCBs (amlodipine, nifedipine, diltiazem, verapamil) aggravated thoracic aortic diameters in Ang II mouse model. **a**, Experiment design. 16-week-old C57BL/6J mice were infused with AngII (1000 ng/kg·min^-1^) for 28 days. After 7 days of infusion, the mice were daily injected with vehicle, dihydropyridine CCBs [amlodipine (1 mg/kg/day) or nifedipine (20 mg/kg/day)], benzothiazepine CCB [diltiazem (8 mg/kg/day)], phenylalkylamine CCBs [verapamil (12 mg/kg/day)]. Groups: Saline + vehicle (n=7); AngII + vehicle (n=13); AngII + amlodipine (n=12); AngII + nifedipine (n=12); AngII + diltiazem (n=11); AngII + verapamil (n=11). **b,** The SBP of mice were recorded by tail-cuff blood pressure measurement at -7, 0, 3,7,14,21 days after vehicle/CCBs treatment. * *vs.* AngII + Vehicle group. **c-e,** Ultrasound-monitored maximal inner diameters of the aortic root, ascending aorta, and aortic arch in mice at different time points. * *vs.* AngII + Vehicle group. **f,** Representative ultrasound images of thoracic aorta after 21 days of vehicle/CCB treatment **g-i,** Monitoring of rd-PWV and ascending aortic strain in mice at different time points by M-mode ultrasound. (g) Schematic diagrams illustrating the calculation. (h) Statistical analysis on rd-PWV. (i) Statistical analysis on ascending aortic strain. * *vs.* AngII + Vehicle group. **j,** Representative *ex vivo* morphology of thoracic aortas from mice. Scale bar=2 mm. **k-m,** Maximal external diameters of ascending aorta, aortic arch and descending aorta. * *vs.* AngII + Vehicle group. b-e, h-i, k-m, Data are the mean ± SEM of each group. \**P* <L0.05; b-e, h-i, Two-way ANOVA with Tukey multiple comparisons test. k-m, Brown-Forsythe and Welch ANOVA test with Dunnett T3 multiple comparisons. AngII, angiotensin II; SBP, systolic blood pressure; CCB, calcium channel blocker; rd-PWV, root to descending aorta pulse wave velocity.

During CCB administration, we performed weekly ultrasound to monitor the diameters of the thoracic aorta (aortic root, ascending aorta, and aortic arch). Compared with those in AngII+Vehicle group, the mice in the CCB groups exhibited significant aortic expansion after 14 days of treatment, which persisted for 21 days. The AngII+verapamil group presented the largest thoracic aortic diameter among the CCB groups (AngII+verapamil *vs.* AngII+vehicle; Aortic root, 2.72±0.10 *vs.* 2.18±0.04 mm, *P*=0.006; Ascending aorta, 2.25±0.12 *vs.* 1.76±0.02 mm, *P*=0.043; Aortic arch, 1.93±0.05 *vs.* 1.64±0.02 mm, *P*=0.006) (Figure 2c-f). Additionally, considering the potential impact of VSMC contractility changes on vascular mechanical properties^15,19,29^, we measured the thoracic root-to-descending pulse wave velocity (rd-PWV) and aortic strain in mice using M-mode ultrasound. These two indicators jointly reflect vascular stiffness and elasticity (Figure 2g). Interestingly, compared with the AngII+Vehicle group, CCB treatment significantly increased the rd-PWV and decreased the ascending aortic strain after 7 days, with these differences becoming more pronounced after 14 and 21 days (Figure 2h-i). These data indicate that CCB treatment increased aortic stiffness in AngII-infused mice, preceding aortic dilation. After 21 days of CCB treatment, the mice were sacrificed, and no significant differences in body weight or plasma lipid levels were observed among the groups (Extended Data Table 13). *Ex vivo* measurements revealed that both amlodipine and nifedipine significantly increased the maximum diameter of the descending aorta compared with that in AngII+Vehicle group. While the AngII+diltiazem and AngII+verapamil groups tended toward larger aortic root diameters, but this difference did not reach statistical significance compared with AngII+Vehicle group. All four CCBs significantly increased the diameters of the ascending aorta and aortic arch (Figure 2j-m). Further Elastic Van Gieson (EVG) staining revealed that CCBs significantly exacerbated elastic fiber breakage and degradation in thoracic aortas (Extended Data Figure 1a). Taken together, these results suggested that all types of CCBs exacerbated TAA (thoracic aortic aneurysm) expansion in AngII-infused mice.

Meanwhile, ultrasound monitoring revealed a decrease in abdominal aortic strain as early as 7 days after CCB treatment, indicating increased vascular stiffness compared to the AngII+Vehicle group (Figure 3a-b). After 14 days of CCB treatment, significant diameter expansion was observed in the suprarenal abdominal aorta, with the AngII+verapamil group showing the largest diameter (Figure 3c). Finally, CCB treatment not only increased the maximum abdominal aortic diameter and the incidence of AAA (AngII+Vehicle, 0% [0/13]; AngII+amlodipine, 41.7% [5/12]; AngII+nifedipine, 41.7% [5/12]; AngII+diltiazem, 45.5% [5/11]; AngII+verapamil, 63.6% [7/11]) (Figure 3d-f), but also consistently exacerbated elastic fiber degradation in the abdominal aorta (Extended Data Figure 1b). Moreover, in an elastase-induced infrarenal AAA model, compared with the Elastase+Vehicle group, CCB treatment further increased the maximum aortic diameter and exacerbated elastic fiber degradation with no significant differences in SBP or body weight among the groups (Figure 4a-f and Extended Data Table 14). Together, these results indicate that CCBs increased the risk of AAA occurrence and development in mice.

**Figure 3.**
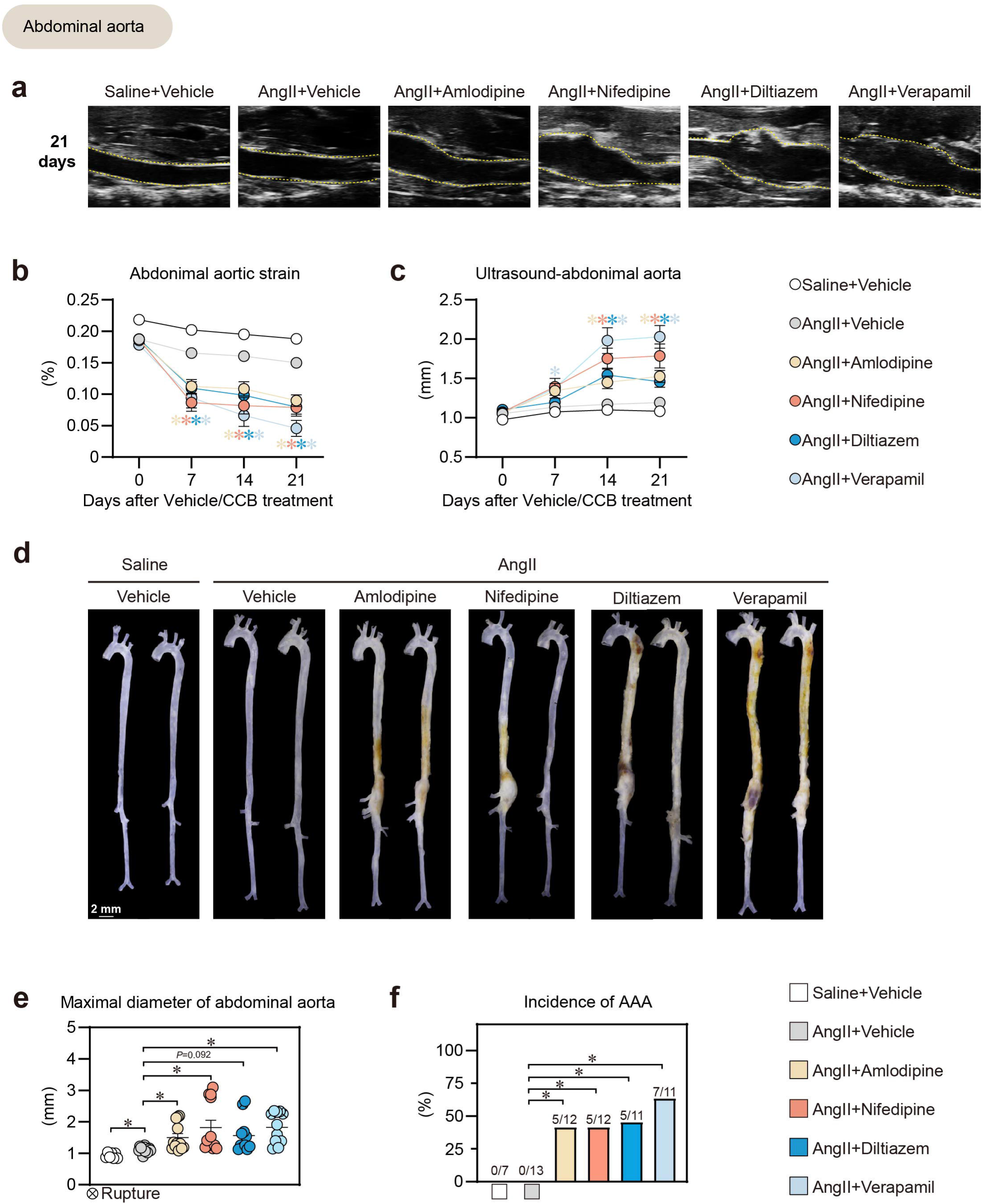
CCBs (amlodipine, nifedipine, diltiazem, verapamil) increased abdominal aortic aneurysm formation in Ang II mouse model. **a**, Representative ultrasound images of abdominal aorta after 21 days of vehicle/CCB treatment. **b-c,** Ultrasound-monitored (b) abdominal aortic strain and (c) maximal inner diameters of abdominal aorta. * *vs.* AngII + Vehicle group. **d,** Representative *ex vivo* morphology of aortas from mice. Scale bar=2 mm. **e-f,** Maximal diameter of (k) abdominal aorta and (l) incidence of AAA. b-c, e, Data are the mean ± SEM of each group. f, Data are presented as percentage (%). \**P* <L0.05; ns, not significant. b-c, Two-way ANOVA with Tukey multiple comparisons test. e, Brown-Forsythe and Welch ANOVA with Dunnett T3 multiple comparisons test. f, Fisher’s exact test. CCB, calcium channel blocker; AngII, angiotensin II. AAA, abdominal aortic aneurysm.

**Figure 4.**
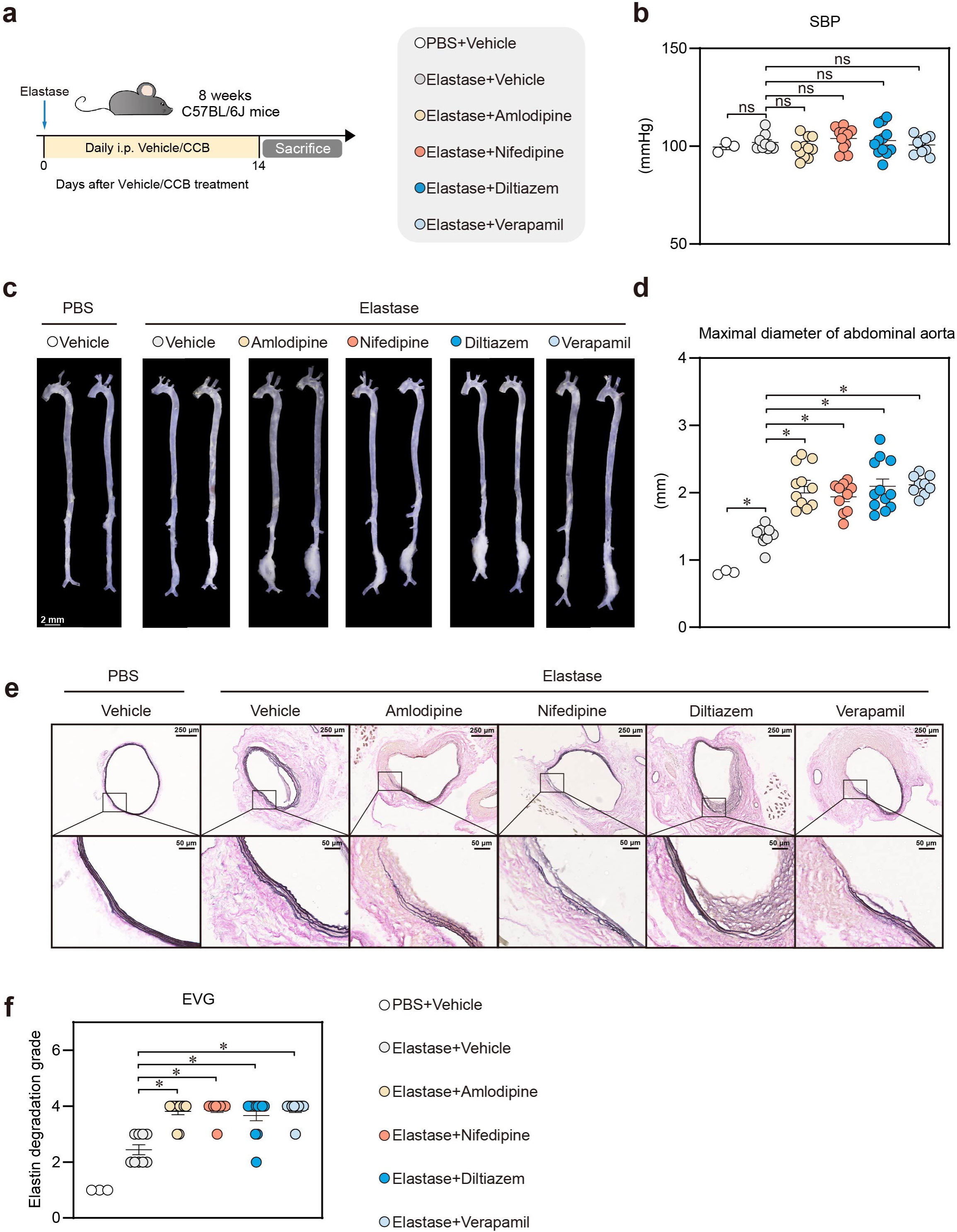
CCBs (amlodipine, nifedipine, diltiazem, verapamil) exacerbated elastase-induced AAA and elastin degradation in mice. **a**, Experiment design. 8-week-old mice infrarenal abdominal aortas were incubated with PBS or elastase (10 mg/ml) and then daily injected with vehicle, amlodipine (1 mg/kg/day), nifedipine (20 mg/kg/day), diltiazem (8 mg/kg/day) or verapamil (12 mg/kg/day) for 14 days. Groups: PBS + vehicle (n=3); Elastase + vehicle (n=9); Elastase + amlodipine (n=11); Elastase + nifedipine (n=11); Elastase + diltiazem (n=12); Elastase + verapamil (n=9). **b,** The SBP of mice were recorded by tail-cuff blood pressure measurement at 14 days after vehicle/CCB treatment. **c,** Representative *ex vivo* morphology of aortas from mice. Scale bar=2 mm. **d,** Maximal diameter of abdominal aorta. **e-f,** Representative of abdominal aorta elastin Van Gieson staining. Scale bar=250/50 μm. b, d, f, Data are the mean ± SEM of each group. \**P* <L0.05; ns, not significant. b, One-way ANOVA with Dunnett multiple comparisons test. d, Brown-Forsythe and Welch ANOVA test with Dunnett T3 multiple comparisons. f, Kruskal-Walli’s test comparisons test. CCB, calcium channel blocker; PBS, phosphate buffered saline; SBP: systolic blood pressure.

### CCBs aggravated BAPN-induced TAAD in mice

Next, we applied a BAPN-induced mouse model of TAAD. Three-week-old C57BL/6J male mice were administered freshly prepared BAPN solution in the drinking water at a dosage of 0.5 g/kg per day for 28 days. After 7 days of BAPN treatment, the mice were injected daily with either vehicle or one of four CCBs. The control mice received normal drinking water (Figure 5a). There were no observable differences in body weight and water intake among the BAPN groups (Extended Data Table 15). As a result, BAPN treatment led to rupture and premature death in 13.3% of the mice. Amlodipine, nifedipine, and verapamil increased the rupture and death rates, whereas diltiazem did not significantly affect mortality (Figure 5b). Compared with the BAPN + Vehicle group, all four CCBs significantly increased the incidence of TAAD (BAPN+Vehicle, 46.7% [7/15]; BAPN+amlodipine, 100% [10/10]; BAPN+nifedipine, 100% [7/7]; BAPN+diltiazem, 85.7% [6/7]; BAPN+verapamil, 90% [9/10]) (Figure 5c-d). *Ex vivo* measurements revealed significant expansion of the ascending aorta, aortic arch, and descending aorta in all CCB-treated groups compared with that in BAPN+Vehicle group (Extended Data Figure 2a-c). EVG staining further revealed that CCBs markedly exacerbated elastic fiber breakage and degradation (Figure 5e and Extended Data Figure 2d). In a parallel experiment, early changes in thoracic aortic mechanical properties were detected using ultrasound (Extended Data Figure 2e), which revealed a significant increase in rd-PWV and decrease in ascending aortic strain after as early as 3 days after the start of amlodipine and verapamil treatment. These effects occurred before any observable aortic dilation and intensified after 7 days (Figure 5f-g and Extended Data Figure 2f-j). Thus, CCBs aggravated BAPN-induced TAAD and aorta stiffness in mice.

**Figure 5.**
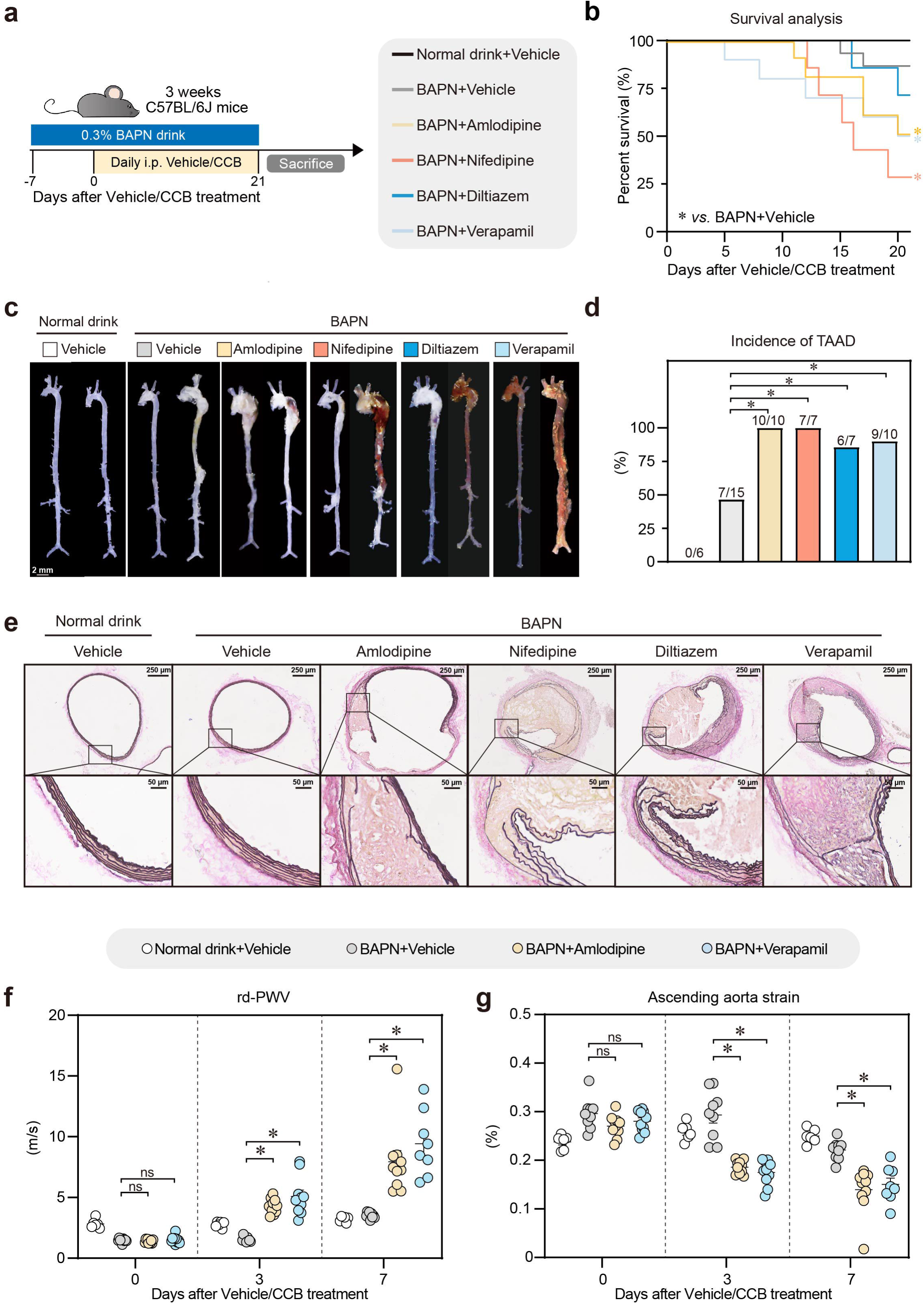
CCBs (amlodipine, nifedipine, diltiazem, verapamil) aggravated BAPN-induced TAAD in mice. **a**, Experiment design. 3-week-old mice were treated with 0.3% BAPN in drinking water (0.5 g/kg per day) for 28 days. Normal drink served as negative control. After 7 days of BAPN treatment, mice were daily injected with vehicle or different CCBs: amlodipine (1 mg/kg/day), nifedipine (20 mg/kg/day), diltiazem (8 mg/kg/day), verapamil (12 mg/kg/day). Groups: Normal drink + vehicle (n=6); BAPN + vehicle (n=15); BAPN + amlodipine (n=10); BAPN + nifedipine (n=7); BAPN + diltiazem (n=7); BAPN + verapamil (n=10). **b,** Survival analysis of mice after vehicle/CCB treatment. *, *vs.* BAPN + Vehicle group. **c,** Representative *ex vivo* morphology of the aortas. Scale bar =2 mm. **d,** Incidence of TAAD. **e,** Representative images of elastin Van Gieson staining. Scale bar = 250/50 μm as shown in the image. **f-g,** Three-week-old mice were treated with 0.3% BAPN in drinking water (0.5 g/kg per day) for 7 days. Then, they were daily injected with vehicle, amlodipine (1 mg/kg/d) or verapamil (12 mg/kg/d) for another 7 days. Rd-PWV (f) and ascending aorta strain (g) were measured by ultrasound at different time points. b, d, Data are presented as percentage (%). f-g, Data are the mean ± SEM of each group. \**P* <L0.05; ns, not significant. b, Survival curve by Logrank test. d, Fisher’s exact test. f-g, Two-way ANOVA with Dunnett’s multiple comparisons test. TAAD, thoracic aortic aneurysm and dissection; CCB, calcium channel blocker; Rd-PWV, root to descending aorta pulse wave velocity; BAPN, β-aminopropionitrile.

### CCBs application limited aortic regression in patients with Type B Aortic Dissection Patients following endovascular therapy

Enabled by the GUEST study (NCT04765605), a single-arm, prospective, multicenter clinical trial that primarily evaluated the treatment efficacy of a novel stent graft system for type B aortic dissection (TBAD) (Figure 6a), we further conducted a secondary analysis to assess the impact of CCB use on aortic expansion. In accordance with the study design, participants underwent computed tomography angiography (CTA) at 1 month, 6 months, and annually after thoracic endovascular aortic repair (TEVAR). Among the 120 enrolled participants, 95 were included in the final analysis, with 69 using CCBs, either exclusively or in combination with other antihypertensive medications and 26 using other antihypertensive medication (Figure 6b and Extended Data Table 16). Baseline characteristics, preoperative laboratory results, and initial aortic morphology are shown in Extended Data Tables 17-18. The alterations in aortic diameters, including those in Ishimaru zone 3 and zone 4, and the maximum descending thoracic aorta diameter (DTA max), as well as volumes in zones 3-4, zone 5, and zones 3-5, were measured and analyzed in strict accordance with the established reporting standards^30^ (Figure 6c and Extended Data Figure 3a).

**Figure 6.**
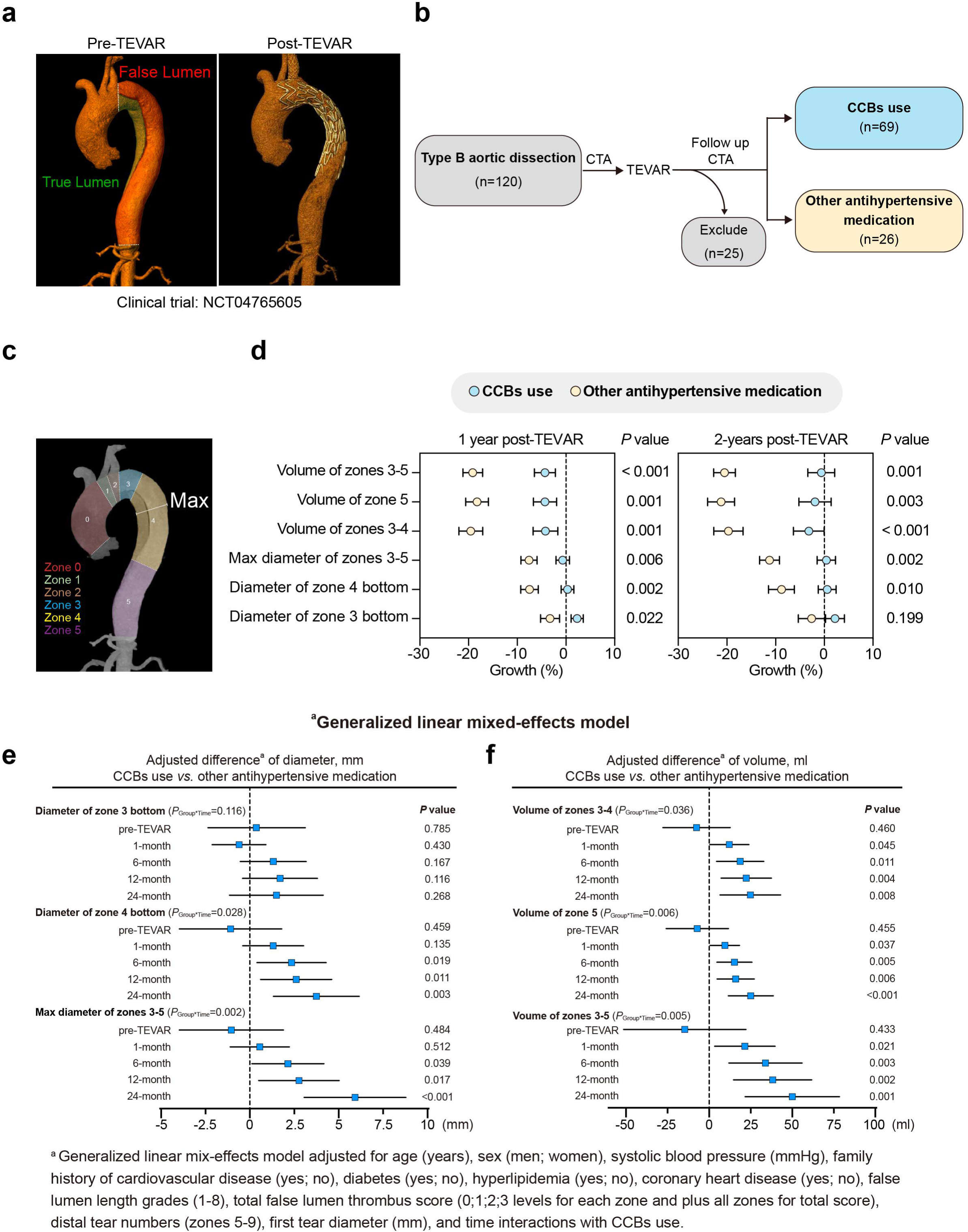
The application of CCBs limited aortic regression in patients with Type B Aortic Dissection Patients following endovascular therapy. **a**, The representative computed tomography angiography (CTA) images of the thoracic aorta in patients with TBAD, showing the condition before (left) and after (right) the application of the covered stent graft system-Weflow-Tbranch^TM^ (a single-embedded branch thoracic aortic covered stent graft system) for endovascular repair. **b,** Patient screening flowchart. **c,** The representative CTA images of a patient before TEVAR surgery show the segmentation of the aorta. Zone 3 (2 cm distal to the left subclavian artery, shown in blue), zone 4 (from the left subclavian artery to the mid - point of the descending thoracic aorta, shown in yellow), and zone 5 (from the mid - point of the DTA to the superior border of the celiac artery, shown in purple). **d,** The growth rates of aortic diameter/volume of patients one and two years after TEVAR surgery. Growth rate (%) = (post-diameter/volume - pre-diameter/volume) / pre - diameter/volume. **e-f,** Adjusted difference of (e) aortic diameters and (f) volumes at pre-TEVAR and 1, 6, 12, 24 months after TEVAR (CCBs use *vs.* other antihypertensive medication). *P* values were present in the figure by generalized linear mix-effects model adjusted for age (years), sex (men; women), systolic blood pressure (mmHg), family history of cardiovascular disease (yes; no), diabetes (yes; no), hyperlipidemia (yes; no), coronary heart disease (yes; no), false lumen length grades (1-8), total false lumen thrombus score (0;1;2;3 levels for each zone and plus all zones for total score), distal tear numbers (zones 5-9), first tear diameter (mm), and time interactions with CCBs. d, Data are the mean ± SEM of each group; e, Data are presented as 95% Cl. d, *P* value < 0.05 except diameter of zone 3 bottom in 2-year after TEVAR by Student’s T test. e-f, *P* values were present in the figure by generalized linear mix-effects model. TEVAR, thoracic endovascular aortic repair. TBAD, type B aortic dissection. CCB, calcium channel blocker. CTA, computed tomography angiography.

As a result, at the 1-year follow-up, the growth rates (%) for all indicators—including zone 3, zone 4, and DTA max diameters, as well as zones 3-4, zone 5, and zones 3-5 volumes—were significantly higher in CCB users compared to those on other antihypertensive medication. By the 2-year follow-up, these elevated growth rates remained significant for all indicators except for the zone 3 diameter (Figure 6d). Using mixed-effects models, we observed that CCB users had greater increases in DTA max zone 4, and zone 3 diameters, as well as larger zones 3-5, zone 5, and zones 3-4 volumes, compared with those in patients using other antihypertensive medications (*P*<0.05 for all, Extended Data Figure 3b-g). After further adjustment for variables such as the aortic dissection phase, false lumen extension, thrombus status in the false lumen, intimal tear number and size, and initial aortic diameter and volume, differences in diameter between the two groups remained significant (zone 4: *P*=0.028, DTA max: *P*=0.002) and became more pronounced over time. Similarly, the adjusted differences in zones 3-4 (*P*=0.036), zone 5 (*P*=0.006), and zones 3-5 (*P*=0.005) volumes were also significant and increased throughout the 2-year period (Figure 6e-f). These findings suggest that TBAD patients who undergo TEVAR and subsequently use CCBs may experience worsened aortic regression/shrinkage.

### PRKG1 silencing alleviates CCB-induced exacerbation of AAD in mice

VSMC contractility is driven primarily by the phosphorylation of the myosin regulatory light chain (p-MLC), which is positively regulated by the intracellular Ca^2+^/calmodulin-myosin light chain kinase (MLCK) pathway. In contrast, protein kinase G type 1 (PRKG1) enhances the activity of myosin light chain phosphatase (MLCP) by inhibiting myosin phosphatase target subunit 1 (MYPT1) phosphorylation, thereby counteracting the phosphorylation of MLC (Figure 7a)^18,31^. Notably, a gain-of-function mutation in PRKG1 has been associated with spontaneous AAD in both patients and transgenic mice^32,33^. We did observe a significantly decrease in p-MLC and p-MYPT1 levels in the aortas of verapamil-treated mice, in both AngII-induced and BAPN-induced AAD models (Figure 7b-c). Therefore, we hypothesized that genetically inhibiting the *Prkg1* to restore VSMC contractility may mitigate CCB-induced aggravation of AAD.

**Figure 7.**
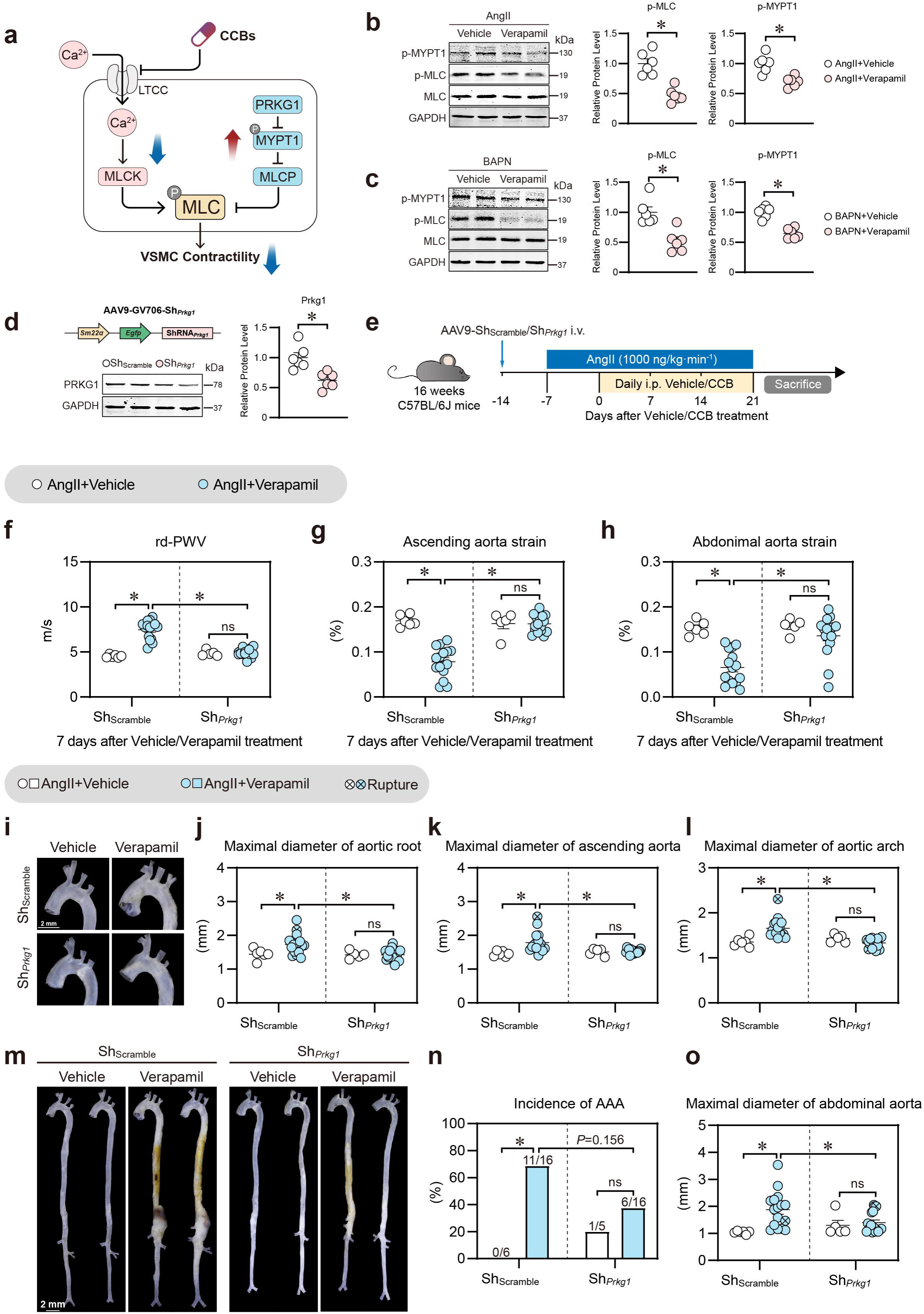
Silencing of *Prkg1* mitigates CCB-aggravated AAD in mice. **a,** Diagram of the signaling involved in vascular smooth muscle cell contraction. **b-c,** Representative western blot image and statistical analysis of MLC phosphorylation, and MYPT1 phosphorylation in aortas from (b) mice infused with AngII (1000 ng/kg·min^-1^) for 7 days or (c) treated with 0.3% BAPN for 3 days followed by injection of vehicle or verapamil (20 mg/kg/d) for 1 hour. **d,** Illustration of the AAV9 vector and statistical analysis of PRKG1 protein level in aorta from mice treated with AAV9-Sh*_Prkg1_* for 2 weeks. **e,** 16-week-old C57BL/6J mice were intravenously injected with AAV9-Sh*_Prkg1_*or AAV9-Sh_Scramble_ for 1 weeks. Then, mice were infused with AngII (1000 ng/kg·min^-1^) for 28 days. After 7 days of AngII infusion, the mice were daily injected with vehicle/verapamil (12 mg/kg/day). Groups: Sh_Scramble_ + AngII + vehicle (n=6); Sh_Scramble_ + AngII + verapamil (n=16); Sh*_Prkg1_*+ AngII + vehicle (n=5); Sh*_Prkg1_*+ AngII + verapamil (n=16). **f-h,** Statistical analysis on ultrasound-monitored (f) rd-PWV, (g) ascending aorta strain, and (h) abdominal aorta strain at 7 days after vehicle/verapamil treatment. **i-o,** (i) Representative *ex vivo* morphology of thoracic aortas. Scale bar=2 mm. (j) Maximal diameters of aortic root. (k) Maximal diameters of ascending aorta. (l) Maximal diameters of aortic arch. (m) Representative *ex vivo* morphology of aortas. Scale bar=2 mm. (n) incidence of AAA. (o) Maximal diameters of abdominal aorta. b-d, f-h, j-l, o Data are the mean ± SEM of each group. n, Data are presented as percentage (%). \**P* <L0.05; ns, not significant. b-d, Unpaired Student T test; f-h, j-l, o, Two-way ANOVA with Tukey multiple comparisons test. n, Fisher’s exact test. LTCC, L-type calcium channel; MLC, myosin regulatory light chain; MLCK, myosin light chain kinase; PRKG1, protein kinase G type 1; MLCP, myosin light chain phosphatase; MYPT1, myosin phosphatase target subunit 1; AAA, abdominal aortic aneurysm; TAAD, thoracic aortic aneurysm and dissection; Rd-PWV, root-descending pulse wave velocity.

An adeno-associated virus 9 (AAV9) vector containing *SM22*α promoter was designed to specifically knockdown *Prkg1* in VSMCs (Figure 7d). Sixteen-week-old C57BL/6J mice were intravenously injected with either AAV9-Sh*_Prkg1_* or AAV9-Sh_Scramble_. After 7 days, the mice received subcutaneous infusions of AngII for 28 days, followed by daily intraperitoneal injections of either vehicle or verapamil starting 7 days after AngII treatment (Figure 7e). Ultrasound results revealed that verapamil significantly increased aortic stiffness in AAV9-Sh_Scramble_-treated mice, but the differences were not observed in the AAV9-Sh*_Prkg1_*-treated mice (Figure 7f-h). As expected, in AAV9-Sh_Scramble_-treated mice, verapamil significantly increased the maximum diameter of the thoracic aorta (including the aortic root, ascending aorta, and aortic arch) (Figure 7i-l), the incidence of AAA (Figure 7m-n), the maximum diameter of the abdominal aorta (Figure 7o) and elastic fiber degradation (Extended Data Figure 4a-d) compared with those associated with vehicle treatment. However, these differences were no longer present in the AAV9-Sh*_Prkg1_*-treated mice. Similarly, in AAV9-Sh*_Prkg1_*-or AAV9-Sh_Scramble_-treated mice subjected to BAPN treatment, verapamil significantly increased the incidence of TAAD in the AAV9-Sh_Scramble_ group (Sh_Scramble_+Vehicle: 45% [9/20]; Sh_Scramble_+Verapamil: 81.25% [13/16]), whereas *Prkg1* knockdown abolished these effects (Sh*_Prkg1_*+Vehicle: 50% [5/10]; Sh*_Prkg1_*+Verapamil: 40% [4/10]) (Extended Data Figure 5a-c). Additionally, *ex vivo* measurements (Extended Data Figure 5d-f) and EVG staining (Extended Data Figure 5g-h) revealed that *Prkg1* knockdown mitigated CCB-induced aortic expansion and elastic fiber degradation, respectively. Taken together, these data indicate that inhibiting the PRKG1-MLCP pathway to restore VSMC contractility can mitigate CCB-aggravated AAD formation.

## Discussion

Antihypertensive drugs are commonly prescribed for AAD due to the established link between hypertension and AAD^13,14^. However, previous studies have primarily focused on the clinical use of antihypertensive drugs in patients with established AAD, while no prior studies have specifically addressed the impact of antihypertensive drugs on AAD incidence among hypertensive patients. Although impaired smooth muscle cell contractility is a known factor in AAD, whether CCBs, which inhibit smooth muscle contraction, increase AAD risk remains unclear. Herein, based on the prospective analysis of 501,878 initially AAD-free participants and 95 TBAD patients who underwent TEVAR, and AAD mouse model, we present an alarming observation that CCBs may increase the risk of AAD and the severity of post-surgical prognosis (Figure 8).

**Figure 8.**
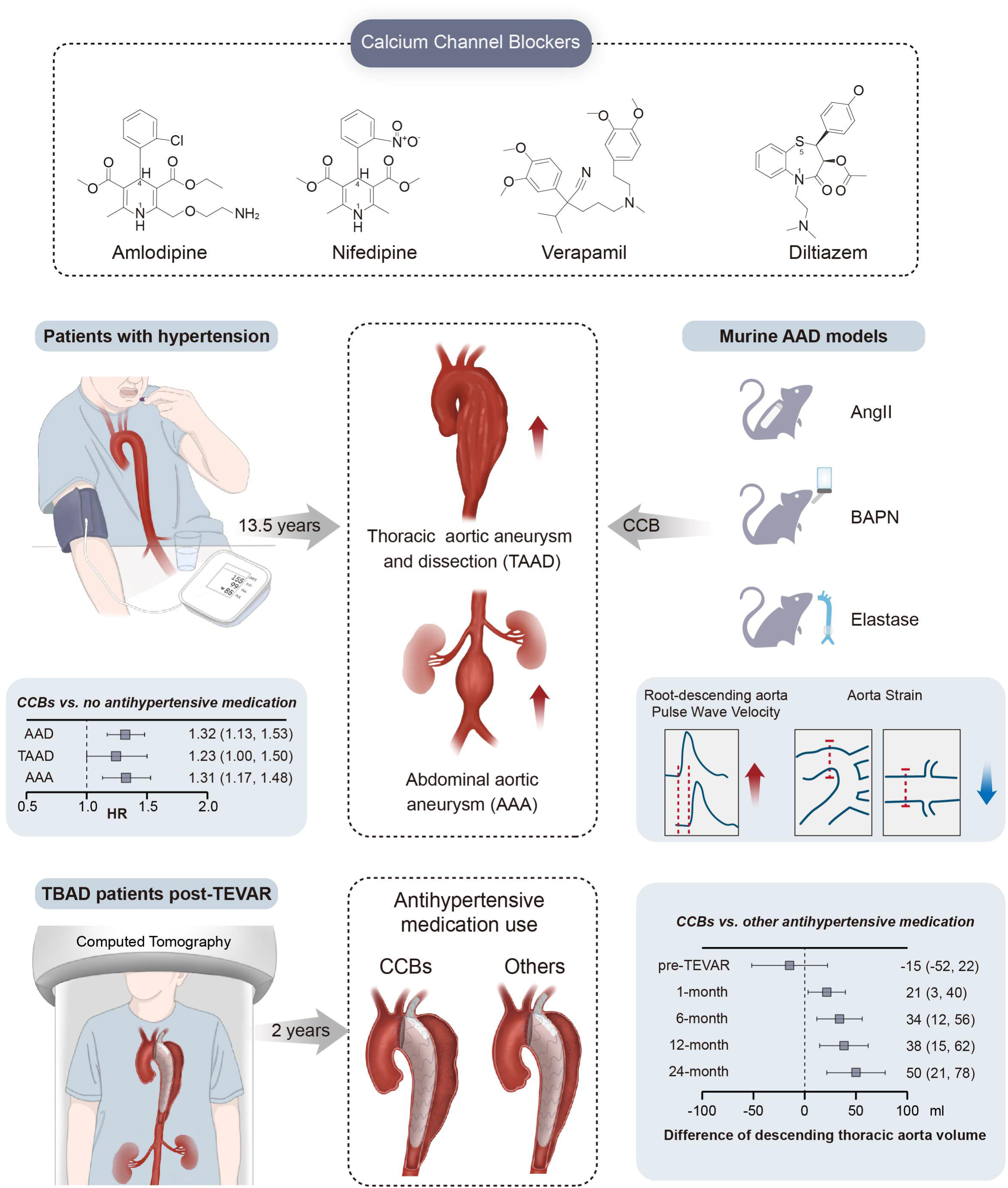
Summary diagram. CCB, calcium channel blocker; AAD, aortic aneurysm and dissection; TAAD, thoracic aortic aneurysm and dissection; AAA, abdominal aortic aneurysm; TBAD, type B aortic dissection; TEVAR, thoracic aorta endovascular repair.

For AAD-related pharmacological studies, currently, lipid-modifying therapies^8^, anti-inflammatory agents^9,10^, antithrombotics^12^, and antibiotics^11,34^ have been evaluated exclusively in AAA, with no evidence of efficacy in limiting aneurysm growth. For antihypertensives treatment (Extended Data Figure 6) in AAD, there are many different types and stages of AAD. Hypertensive patient’s course in the context of AAD can be divided into three stages: the pre-AAD stage (hypertension without AAD), the diagnosed AAD stage, and the post-surgical stage following AAD repair. Since most studies have focused on the treatment of patients already diagnosed with AAD, this stage has the most abundant evidence, particularly in thoracic aortic aneurysm (TAA) research represented by Marfan syndrome. Both the 2022 American Heart Association (AHA)^35^ and 2024 European Society of Cardiology (ESC)^36^ recommend the use of ARBs and β-blockers in patients with MFS due to their reducing the rate of progressive aortic root enlargement^37–39^. In cases of acute aortic dissection, β-blockers or CCB are recommended as the first-line therapy to reduce blood pressure and heart rate, thereby minimizing further dissection. Beyond these scenarios, however, for non-genetic forms of TAAD and AAA, evidence remains limited— only two randomized controlled trial in patients with small AAAs found no effect of amlodipine or telmisartan on AAA^5,7^. Of note, no studies have evaluated whether the use of antihypertensive medications influences the risk of AAD incidence in hypertensive patients.

Herein, by analyzing data from 501,878 initially AAD-free participants from the UKB, we found that CCB use in individuals with hypertension was associated with a significantly increased risk of new-onset AAD, with phenylalkylamine CCBs showing the highest risk. The main advantage of UKB analysis lies in its use of large-scale population data, overcoming the challenge of conducting randomized controlled trials for AAD due to its low incidence. Additionally, this analysis leveraged a prospective design from the UKB. These elements enabled a detailed assessment of the association between CCB use and AAD, with adjustments for potential confounders. The concurrent use of other antihypertensives was also considered to minimize indication bias, supporting the observed positive association between CCB use and AAD. Furthermore, we found that the administration of all three CCB subtypes—dihydropyridines, phenylalkylamines, and benzothiazepines—increased the risk of AAD in AngII-, elastase-, and BAPN-induced mouse models. Consistently, a previous study has shown that CCB increased aorta expansion in MFS^27^. Of interest, several studies reported a potential protective effect of amlodipine and diltiazem against AngII-induced AAD in hyperlipidemic mice^24,25^. However, the doses used in those studies were 5-10 times higher than those in our study or clinical practice. Furthermore, previous studies exclusively applied concurrent CCBs and AngII on the establishment of the hypertension model^23,26^. In our study, to mimic the *bona fide* situation of hypertensive populations, we implanted AngII pumps in 4-month-old wild-type mice for one week to establish hypertension prior to administering CCB drugs. Regarding the association between postoperative antihypertensive use and outcomes in patients with AAD after surgical repair, only a few studies reported. One study reported that CCB use was associated with a 3.41-fold increase in perioperative mortality after open surgical repair of AAD^40^, while another suggested that β-blocker therapy may reduce long-term mortality following surgery for TBAD^41^. However, these studies varied significantly in surgical approaches (endovascular and open repair), techniques, and materials used, all of which can greatly influence postoperative outcomes. In our study, all patients had TBAD and underwent TEVAR using the same stent graft system and standardized implantation protocol. The secondary retrospective analysis revealed that CCB use in these patients was associated with worsened aortic regression. These results highlight the importance of careful antihypertensive selection in this population and emphasize the urgent need for safer, more effective treatment strategies in AAD prevention and post-surgery management.

Another intriguing finding of the current study is that impaired VSMC contractility is a key factor in the exacerbation of AAD induced by CCBs. Previous research has linked mutations in essential contractile genes (*MYH11* and *ACTA2*)^42,43^ and regulators such as *MLCK*^44^ and *PRKG1*^33^ to AAD formation. Additionally, several drugs that may impair VSMC contractility, such as sildenafil and fluvastatin, have been shown to aggravate aneurysm progression^45,46^. However, the exact mechanisms by which VSMC contraction dysfunction contributes to AAD remain unclear. Herein, data from a natural population cohort, a prospective post-AAD cohort, and animal models consistently demonstrated that CCB use, which inhibits VSMC contractility, significantly increased the risk of AAD. This raises the following question: why does CCB-induced inhibition of VSMC contractility worsen AAD? We found that CCB treatment in AAD model mice significantly suppressed VSMC contractility, as characterized by decreased MLC and MYPT-1 phosphorylation. In contrast, *in vivo* knockdown of PRKG1, an endogenous inhibitor of MLC phosphorylation, restored VSMCs contractility and notably mitigated the CCB-induced exacerbation of AAD. This finding aligns with prior findings showing that gain-of-function mutations in PRKG1 lead to spontaneous aortic dissection, whereas PRKG1 knockdown or inhibition reduces aortic dilation in Marfan syndrome (MFS) mice and elastase-induced AAA models^46,47^. Using ultrasound, we observed that in both AngII- and BAPN-induced AAD models, CCB treatment increased rd-PWV and reduced aortic strain, indicating increased aortic stiffness and decreased elasticity. Notably, these mechanical changes occurred before aortic dilation and aneurysm formation, consistent with earlier studies showing increased aortic stiffness in AAA patients treated with CCBs and elevated VSMC stiffness in tissues from TAAD or AAA patients and animal models^19,29,48,49^. Our findings underscore the critical role of VSMC contractility in AAD progression and suggest that ultrasound monitoring of vascular mechanics may serve as an effective strategy for early warning, prediction and intervention of AAD. Whether other drugs affecting contractility, such as nitrates, potassium channel openers, and Rho kinase inhibitors, may also exacerbate AAD and warrant further investigation.

The limitations of this study include the lack of detailed data on the frequency, duration, and dosage of CCB use, although self-reported medication information was recorded and verified by trained nurses during the same visit. The participants of the UKB are predominantly White British, well-educated, and affluent participant base, with lower rates of smoking and drinking, which may limit the generalizability of our findings to other populations. However, similar detrimental effects of CCBs were observed in Chinese patients with TBAD after stent implantation, suggesting the robustness of our results. Although misclassification in CCB use or AAD outcomes is possible, it is likely nondifferential due to the prospective study design. Finally, due to the small size of BAPN-treated mice, physical restrictions leading to potential TAAD rupture limited our ability to obtain blood pressure data.

In summary, our findings highlight a concerning observation, indicating the detrimental impact of CCBs on the risk of AAD.

## Sources of Funding

This research was supported by funding from the National Key R&D Program of China (2022YFA1302900 to W. K., 2024YFC2419000 to W. G.), the National Natural Science Foundation of China (NSFC 82230010 to W.K., 82070494 to W. G., 82473622 to X. G., and 82300544 to Z.Y. C.), Science and Technology Project of Tianjin Municipal Health Commission (TJWJ2023QN116 to L. C.)

W.K.; W.G.; X.G. Correspondence and requests for materials should be addressed to Wei Kong.

## Author contributions

W.K., W.G., X.G., Z.Y.C conceptualized and designed the study. W.K., W.G., X.G., Z.Y.C supervised the study. T.F.M., Z.Y.C., X.M.X., L.C. and A.W. performed the experiments and analyzed the data. T.F.M., Z.Y.C., X.M.X. and L.C. wrote the manuscript with input from all authors. T.F.M., Z.Y.C., X.M.X. and L.C. contributed equally to this work. All authors edited the manuscript and approved the final manuscript.

## Competing interest declaration

The authors have declared that no conflicts of interest exist.

## Data Availability

All data produced in the present work are contained in the manuscript

## Materials and Methods

### UKB study population

The UK Biobank (UKB) is a large population-based cohort, including over 500,000 volunteers aged between 40 and 69 years across England, Scotland, and Wales recruited between 2006 and 2010 (Details could be found at https://www.ukbiobank.ac.uk). All UKB participants provided informed consent via an electronic signature at the beginning of the process and the UKB had ethical approval from the North West Multi-Center Research Ethics Committee (REC reference: 16/NW/0274). Included were 501,878 participants in this study after removing participants who withdrew from the UKB and prevalent aortic aneurysm and dissection (AAD). We also investigated the association between subtypes of CCBs and AAD in the participants with prevalent hypertension (n=142,419) and the association between the CCBs use and all-cause mortality in the participants with prevalent aortic aneurysm and dissection (n=509).

### Assessment of exposure

Baseline hypertension was defined through self-reported hypertension during nurse interview, and self-reported questionnaires regarding high blood pressure diagnosed by doctor, hospital data via international classification of diseases (ICD) codes for essential (primary) hypertension, hypertensive disease with or without heart failure, hypertensive heart and renal diseases, and secondary hypertension according to previous study^50^ (Extended Data Table 1). A high concordance was observed between self-reported (either self-reported hypertension during nurse interview or self-reported high blood pressure diagnosed by doctor) and hypertension identified through hospital records, with more than 94% of individuals who self-reported hypertension also having a physician-confirmed diagnosis based on ICD codes. We also adopted a broader definition of hypertension by further incorporating antihypertensive medications use in the sensitivity analysis. Antihypertensive medications regarding calcium channel blocker use (Extended Data Table 2) and other blood pressure lowering medications including renin-angiotensin system inhibitors, beta blockers, diuretic, or multiple antihypertensive classes were assessed at baseline^50,51^ (Extended Data Table 3). All self-reported medications were recorded and subsequently confirmed by a trained nurse in an interview conducted during the same visit. Additionally, participants who reported taking a blood pressure medication (via Field IDs 6153 and 6177) without using CCBs were categorized under the “other antihypertensive medications” group (excluding CCB use). Anyone reporting concurrent CCB use was categorized into the CCBs group, including those using CCBs exclusively or in combination with other antihypertensive medications. There was no recorded information concerning the dosage, frequency, or duration of use for medication. We categorized groups according to hypertension status and antihypertension medications (Figure 1).

### Assessment of covariates

At baseline, data on sociodemographic factors (age, sex, education, and ethnicity), BMI, and lifestyle factors (smoking status and alcohol use) were collected through questionnaires and verbal interviews. Total cholesterol, total triglycerides, and systolic blood pressure were also analyzed in this study, regarding to the risk of AAD. Specifically, serum lipid levels were measured by biochemical assays from the blood samples collected at baseline by Beckman Coulter AU5800 Platform. Total cholesterol and total triglycerides were analyzed using glycerol-3-phosphate (GPO)-peroxidase (POD) chromogenic method. The resting blood pressure was measured by Omron 705 IT electronic blood pressure monitor. During the blood pressure measurement, the participant was instructed to sit with their feet parallel, toes pointing forward, and the soles of both feet flat on the floor. The right arm was only used if the left was not feasible. The mean value of blood pressure readings would be calculated and used in this analysis if there were more than one measurement within a few minutes available; otherwise, electronic blood pressure monitor values were preferred, and manual readings would be used if there were no electronic blood pressure readings.

### Assessment of outcome

AAD cases and its subtypes including thoracic aortic aneurysm and dissection (TAAD) and abdominal aortic aneurysm (AAA) were identified from the UKB using hospital inpatient records where these conditions were documented as either primary or secondary diagnosis, or linkage to death register data using the International Classification of Disease (ICD) coding 10 (Extended Data Table 4).

### Materials

Amlodipine (HY-B0317), Nifedipine (HY-B0284), Diltiazem (HY-14656), Verapamil (HY-14275), human angiotensin II (HY13948) was purchased from Med Chem Express (Monmouth Junction, NJ, USA). Osmotic pumps (2004W) were purchased from RWD Life Science Co (Shenzhen, Guangdong, China). Elastase, porcine pancreas (HY-P2974) was purchased from Med Chem Express (Monmouth Junction, NJ, USA). β-aminopropionitrile monofumarate (BAPN, A3134) was purchased from Sigma Aldrich (St. Louis, MO, USA). PEG200 (P8490), Tween 80 (T8360) and protein phosphatase inhibitor (P1260) were purchased from Solarbio (Beijing, China). Antibodies against GAPDH (10494), PRKG1 (21646) were purchased from ProteinTech (Chicago, IL, USA). Antibody against p-MLC (3671), MLC (8505), p-MYPT1 (4563) was purchased from Cell signaling Technology (Boston, MA, USA).

### Animals

Animal studies were conducted in compliance with the guidelines of the Institutional Animal Care and Use Committee of Peking University Health Science Center and were approved by the Biomedical Ethics Committee of Peking University. SPF mice of C57BL/6J were purchased from the Experimental Animal Science Center and raised in the Experimental Animal Science Center of Peking University Health Science Center, Beijing, China. The mice were randomly assigned to different groups and housed in cages with 4-6 mice per cage with unrestricted access to rodent feed and water. Specifically, a light-dark cycle was established with lights on from 8 a.m. to 8 p.m., while maintaining a temperature range of 21-24 °C and humidity levels between 40-70%. Mice for surgeries were anesthetized with isoflurane (1.5%-2%). All animals were anesthetized with 1.25% avertin before being sacrificed. For plasma samples, blood was collected from the heart with heparin sodium (250U/ml), followed by centrifugation for 10 min at 3000 rpm. For aorta protein samples, mice were perfused with ice-cold saline, and then the aorta was isolated. For aortic morphology studies, the mice were successively perfused with 10% KCl, 10×PBS, and 4% paraformaldehyde. Then, the aortas were dissected from the root to the iliac bifurcation in all animal models for further analysis.

### CCB drugs dissolution and injection

The doses of CCBs used in the experiment were converted from human to mouse doses based on body surface area. The specific doses were as follows: amlodipine (1 mg/kg/day), nifedipine (20 mg/kg/day), diltiazem (8 mg/kg/day) and verapamil (12 mg/kg/day). The dissolution protocol used a gradient of DMSO, PEG 200, Tween 80, and saline. The final CCB injection solution consisted of 2% DMSO, 8% PEG 200, 1% Tween80, and 89% saline. Experimental mice were administered the specified dose of CCBs or vehicle daily via intraperitoneal injection based on body weight, continuing until the sacrifice endpoint.

### Angiotensin II-induced aortic aneurysm mouse model

Sixteen-week-old C57BL/6J mice were subcutaneously infused with minipumps containing AngII (1000 ng/kg·min^-1^) for 28 days. After 7 days of AngII infusion, images of mice thoracic and abdominal aorta were obtained via ultrasound instrument. Developed aorta aneurysm mice were excluded, and only those without aorta aneurysm were retained. Subsequently, those hypertensive mice without aneurysms were daily injected with vehicle or one of four CCBs: amlodipine, nifedipine, diltiazem or verapamil. SBP was recorded at 0, 3, 7, 14, 21 days after vehicle/CCBs treatment. Root-descending aorta pulse wave velocity, ascending and abdominal aorta strain, thoracic and abdominal aorta diameters were detected at 0, 7, 14, 21 days after vehicle/CCBs treatment by ultrasound.

At the sacrificed endpoint, aortas were dissected for *ex vivo* measurements of the maximal thoracic aorta diameter (aortic root, ascending aorta, aortic arch) and abdominal aorta diameter. Aortic aneurysm was evaluated in the suprarenal abdominal aorta (referred to as AAA) and the aortic root and ascending aorta (referred to as TAA). The maximal diameters of the root, ascending and suprarenal aortas were calculated by statistical analysis, and the aortic aneurysm was defined as an aorta with a diameter more than 1.5 times larger than its adjacent normal aorta.

### Elastase-induced abdominal aorta aneurysm mouse model

Eight-week-old C57BL/6 mice were used for surgery. Briefly, the mice were anesthetized and the infrarenal aortas were exposed, isolated, and wrapped with sterile cotton, which was soaked with 25 μl of elastase (10mg/ml) for 40 minutes. PBS was used as negative control. Then, the elastase-soaked cotton was removed, and 0.9% NaCl was used to perfuse the abdominal cavity before suturing. Subsequently, these mice were administered intraperitoneal injections of vehicle or four CCBs for 14 days.

At sacrificed endpoint, systolic blood pressures were measured and aortas were dissected for *ex vivo* measurements of the maximal abdominal aorta diameter.

### BAPN-induced TAAD mouse model

Three-week-old C57BL/6 mice were treated with 0.3% BAPN in drinking water (0.5 g/kg per day) for 28 days. The drinking water was refreshed every 3 days. Normal drinking was designed as negative control. After 1 week, the mice were daily injected with vehicle or four CCBs. It is important to note that due to the small size of the TAAD model mice and the potential risk of TAAD rupture induced by stress during blood pressure measurement, this model is not suitable for recording SBP.

At the sacrificed endpoint, aortas were dissected for *ex vivo* measurements of the maximal thoracic aorta diameter (ascending aorta, aortic arch and descending aorta). The incidence rate of TAAD and related mortality were recorded. TAAD was defined as thoracic aortic dissection (aortic false lumen formation) and/or thoracic aortic aneurysm (at least 1.5 times extension of aortic diameter). Overall, any of the following criteria was defined as TAAD: 1. The deceased mice displayed blood clotting in the thoracic cavity. 2.EVG staining revealed aortic false lumen formation. 3. > 1.5 times extension of aortic diameter compare to negative control.

In another parallel BAPN experiment, ultrasound was used to measure root-descending aorta pulse wave velocity, ascending aorta strain, thoracic aorta diameters at 0, 3, 7 days after vehicle/amlodipine/verapamil treatment.

### Mouse blood pressure measurement

Mouse blood pressure was measured according to the previous study^52^. Briefly, a noninvasive computerized CODA tail-cuff blood pressure system (Kent Scientific, Torrington, CT, USA) was used to measure mouse SBP. The equipment was maintained in a clean state, free from foreign scents or blood odors. The investigator was blinded to the experimental groups when performing the measurements, and the mice were tested in a randomized order. All mice underwent training sessions from 1 to 4 PM on 7 consecutive days to become accustomed to the tail-cuff procedure.

### Mouse aortic ultrasound

Vevo LAZR system (FUJIFILM VisualSonics, Canada) was used to measure aorta parameters in mice, including thoracic aortic diameter, abdominal aortic diameter, root to descending pulse wave velocity (rd-PWV), ascending aorta and abdominal aorta circumferential strain. Mice were anaesthetized by 1.5-2% isoflurane and placed on the platform that carries electrocardiogram (ECG) electrodes. Conductive gel was applied to the electrodes on the animal platform, and the mouse’s paws were secured onto the electrode pads with tape to ensure accurate ECG acquisition. The mouse’s body temperature and respiratory rate were maintained, with the heart rate kept within the range of 400-550 bpm during the acquisition. MX transducer MX550D (centre transmit: 40 MHz; axial resolution: 40 μm) was used. The thoracic and abdominal aorta diameters were recorded by B-mode ultrasonography. For aorta diameter, images of the aortic arch cross-section (the operation table was tilted 15° to the right, and the probe arm was tilted 30-45° to the left) and abdominal aorta sagittal section (the probe was positioned perpendicular to the animal) were obtained. Rd-PWV was measured using ultrasound imaging by B-mode and Doppler-Mode. The vascular pulse-wave Doppler tracing was recorded at the position of root (position 1, d0). The time from the R point of QRS to the onset of the Doppler waveform was measured (T1). Then another position in the descending aorta was selected and the pulse-wave Doppler tracing was recoded (position 2, d). The time from the R point of QRS to the onset of the Doppler waveform was measured (T2). The distance between position 1 and position 2 was measured (distance). Rd-PWV is calculated as: distance/(T2–T1) (m/s). The values for T1 and T2 were the average of three to five measurements. For ascending aorta and abdominal aorta circumferential strain, the systolic diameter (Ds) and diastolic diameter (Dd) were measured, and circumferential cyclic strain was calculated as (Ds-Dd)/Dd. The values for Ds and Dd were the average of three to five measurements.

### Blood biochemical analysis

Blood samples were collected and underwent centrifugation. The quantification of plasma cholesterol and triglyceride levels was performed using commercially available kits (#100000220, Zhong Sheng Biotechnology).

### Elastin *Van Gieson* staining

Cryosections of the lesion aorta (8 μm thick) were analyzed by elastic *Van Gieson* staining (BASO Precision Optics Ltd., Taiwan, China) according to the manufacturer’s protocol for elastin assessment. Elastin degradation graded as grade 1, < 25% degradation; grade 2, 26% to 50% degradation; grade 3, 51% to 75% degradation; grade 4, > 75% degradation.

### Adeno-associated virus 9 (AAV9) production and infection

A plasmid (GV706) encoding promoter *SM22*α, *EGFP* and shRNA targeting mouse *Prkg1* was engineered by cloning the following shRNA into the AAV9 vector:

Sh*_Prkg1_* (sense) 5’-GATCCCCGGAGAATCTCATCCTAGATCTCGAGATCTAGGATGAGATTCTCCGGTTT TTGC-3’;

Sh*_Prkg1_*(antisense) 5’-GGCCGCAAAAACCGGAGAATCTCATCCTAGATCTCGGATCTAGGATGAGATTCTC CGGG-3’;

The sequence of the mouse *Prkg1* shRNA was referred to the previous study^53^. Packaging and producing work were performed by GENE company (Shanghai, China). For in vivo infection experiments, every mouse was intravenously administered AAV9-Sh_Scramble_ and AAV9-Sh*_Prkg1_* with 2×10^11^v.g. titer. Infection efficiency was analyzed in thoracic aorta samples by western blot.

### Western Blot

Mouse tissues or cells were lysed in RIPA buffer (P0013B, Beyotime, Beijing, China) supplemented with protein phosphatase inhibitors (1:100). Protein concentrations were quantified using a BCA Protein Assay Kit (Thermo Scientific). Equal amounts of total proteins were resolved on 8% or 12% SDS-PAGE gels transferred onto nitrocellulose filter membranes (Pall, Port Washington, NY, USA). The membranes were blocked with 5% BSA and then incubated with the primary antibody and the IRDye 700/800DX-conjugated secondary antibody (Rockland Inc., Gilbertsville, PA, USA). The immunofluorescent signal was obtained using an Odyssey infrared imaging system (LI-COR Biosciences, Lincoln, NE, USA).

### TBAD study and Anatomical characteristics

For patient inclusion and exclusion criteria, we excluded deceased patients for whom CTA data could no longer be obtained and patients with major postoperative complications, such as stroke, respiratory failure, retrograde type A aortic dissection, re-intervention, stent-induced new entry, stent migration, stenosis or occlusion of the LSA branch stent, and spinal cord ischemia, due to their poor health status in subsequent follow-ups. In addition, several patients who were not hypertensive were also excluded from this study.

For antihypertensive medication, we confirmed and recorded the patients’ use of antihypertensive medication through discharge medication records, subsequent follow-up information from outpatient visits at the central hospital, and telephone follow-ups.

For CTA measurements, briefly, a series of morphological parameters of TBAD were measured on the preoperative and postoperative CTA images. The measurement protocol strictly followed the reporting standards for TBAD recommended by the Society for Vascular Surgery and Society of Thoracic Surgeons^54^. All measurements were obtained using post-processing image software (3Mensio Workstation version 10.4, Maastricht, the Netherlands) that enabled aortic centre lumen line extraction and reconstruction. Aortic diameter was measured in three fixed planes, including: (1) 2 cm distal to the left subclavian artery (LSA); (2) the middle point of the descending thoracic aorta (DTA) and (3) the superior border of the celiac artery (CA). Additionally, the maximal diameter of the DTA was also recorded. Suggested by the Society for Vascular Surgery and Society of Thoracic Surgeons, the aortic diameter was measured from the outer aortic wall to outer aortic wall, and the diameters of the true lumen (TL) and false lumen (FL) on these four interested observed planes were simultaneously measured. The volumes of the DTA (Ishimaru zones 3-4 and zone 5) was also measured by using the “volume segmentation function” of the post-processing software 3Mensio Vascular. The descending thoracic aorta is anatomically divided into the Ishimaru zones 3-4 and zone 5 (Extended Data Figure 3a).

### Statistical Analysis

For UKB data, we summarized the baseline characteristics of the study stratified by hypertension and CCBs use or other antihypertensive medication, or hypertension without antihypertensive medication use, or no hypertension (Figure 1 and Extended Data Table 5). Person-time was calculated from the baseline to the occurrence of study outcomes, death, or the end of follow-up (October 31, 2022 [England], July 31, 2021 [Scotland], and February 28, 2018 [Wales]), whichever came first. Cox proportional hazards regression models were applied to examine the risk of AAD or AAD subtypes, adjusting for age, sex, BMI, total cholesterol, triglycerides, smoking status, education, ethnicity, and systolic blood pressure. Sensitivity analyses were conducted to minimize the bias: (1) a broader definition of hypertension was applied; (2) restricted to participants without coronary artery disease to address the issue that coronary artery disease might confound the association between CCBs use and AAD; (3) the Fine & Gray competing risk model to account for the presence of competing events (death from all causes except for AAD). We tested the proportional hazard assumption with Schoenfeld residuals and did not detect any significant violation of this assumption. Among participants with hypertension, mixed-effects linear models with unstructured variance structure were used to analyze the effect of groups (without any antihypertensive medications, CCBs use, and other antihypertensive medications excluding CCBs use) on systolic/diastolic blood pressure, with participants treated as random effects, group, time (baseline or follow-up), and their interaction as fixed effects, adjusting for age, sex, BMI, total cholesterol, triglycerides, smoking status, education, ethnicity, and baseline systolic/diastolic blood pressure. Estimated marginal means and 95% CI were the adjusted values from the models and post hoc pairwise comparisons used Bonferroni correction. Statistical analyses for the UKB were performed using R version 3.6.3.

For experiments data, statistical analyses were performed using GraphPad Prism 8.0 software (San Diego, CA, USA). Data are expressed as mean ± SEM. The details of the statistical analysis used in each experiment are presented in the corresponding figure legends. For statistical comparisons, we first evaluated whether the data were normally distributed using the Shapiro Wilk normality test. For two groups, we used unpaired Student’s *t* test for normally distributed data. If the data was skewed distribution, we used Mann-Whitney test. For more than two groups with one variable, we used one-way ANOVA followed by post hoc analysis for comparisons among normally distributed data. Specifically, Dunnett’s multiple comparison test was used to compare each group with a control group, while Tukey’s multiple comparisons test was used to compare each group with every other group. If the data equal variances were not assumed, Brown-Forsythe and Welch ANOVA test was used with Dunnett T3 multiple comparisons test (compare each group with a control group) or Games-Howell multiple comparisons test (compare each group with every other group). For more than two groups with two variables, we used two-way ANOVA (mixed model) among normally distributed data. Specifically, Dunnett’s multiple comparison test was used to compare one variable within each another variable to control group. If the data equal variances were not assumed, Geisser-Greenhouse correction was used. Nonparametric tests (Kruskal-Walli’s test) were used when the data were not normally distributed. In addition, Fisher’s test was used for contingency data and Log-rank test was used Kaplan-Meier curve.

For human study, statistical analyses were performed using GraphPad Prism 8.0 software (San Diego, CA, USA) and SPSS Statistics 26 (IBM, NY, USA). Data are expressed as mean ± SEM or percentages. For statistical comparisons, unpaired Student’s *t* test and χ test (Fisher’s test) was used for the comparison of patient’s baseline characteristics, CTA measurements and diameters or volumes growth rates at 1- or 2-year post-TEVAR. Mixed-effects model without adjustment was used to compare whether there are statistically significant differences in the trends of diameters and volumes between the two groups over a 2-year follow-up period. This model accounts for repeated measures within subjects and evaluates changes over time without adjusting for potential confounding variables. Furthermore, we applied a general linear mixed-effects model, adjusting for potential influencing factors, including age (years), sex (men; women), systolic blood pressure (mmHg), family history of cardiovascular disease (yes; no), diabetes (yes; no), hyperlipidemia (yes; no), coronary heart disease (yes; no), false lumen length grades (1-8), total false lumen thrombus score (0;1;2;3 levels for each zone and combined score for all zones), number of distal tears (zone 5-9), initial tear diameter (mm), and interactions of these factors with CCB use. We then compared whether the trends of diameters and volumes between the two groups over the 2-year follow-up period showed statistically significant differences. Additionally, we assessed whether the differences in diameter and volume between the two groups at preoperative and postoperative time points (1, 6, 12, and 24 months) were statistically significant.

In all analyses, *P* value < 0.05 was regarded as statistically significant.

## Notes

### Competing Interest Statement

The authors have declared no competing interest.

### Funding Statement

This study was funded by the National Key R&D Program of China (2022YFA1302900 to W. K., 2024YFC2419000 to W. G.), the National Natural Science Foundation of China (NSFC 82230010 to W.K., 82070494 to W. G., 82473622 to X. G., and 82300544 to Z.Y. C.), Science and Technology Project of Tianjin Municipal Health Commission (TJWJ2023QN116 to L. C.)

### Author Declarations

Ethics Committee of North West Multi-Center Research gave ethical approval for this work Ethics Committee of Chinese People's Liberation Army General Hospital gave ethical approval for this work.

