## Extended Data Figure 1-6; Extended Data Table 1-20 for "Calcium Channel Blockers Increased the Risk of Aortic Aneurysm and Dissection"

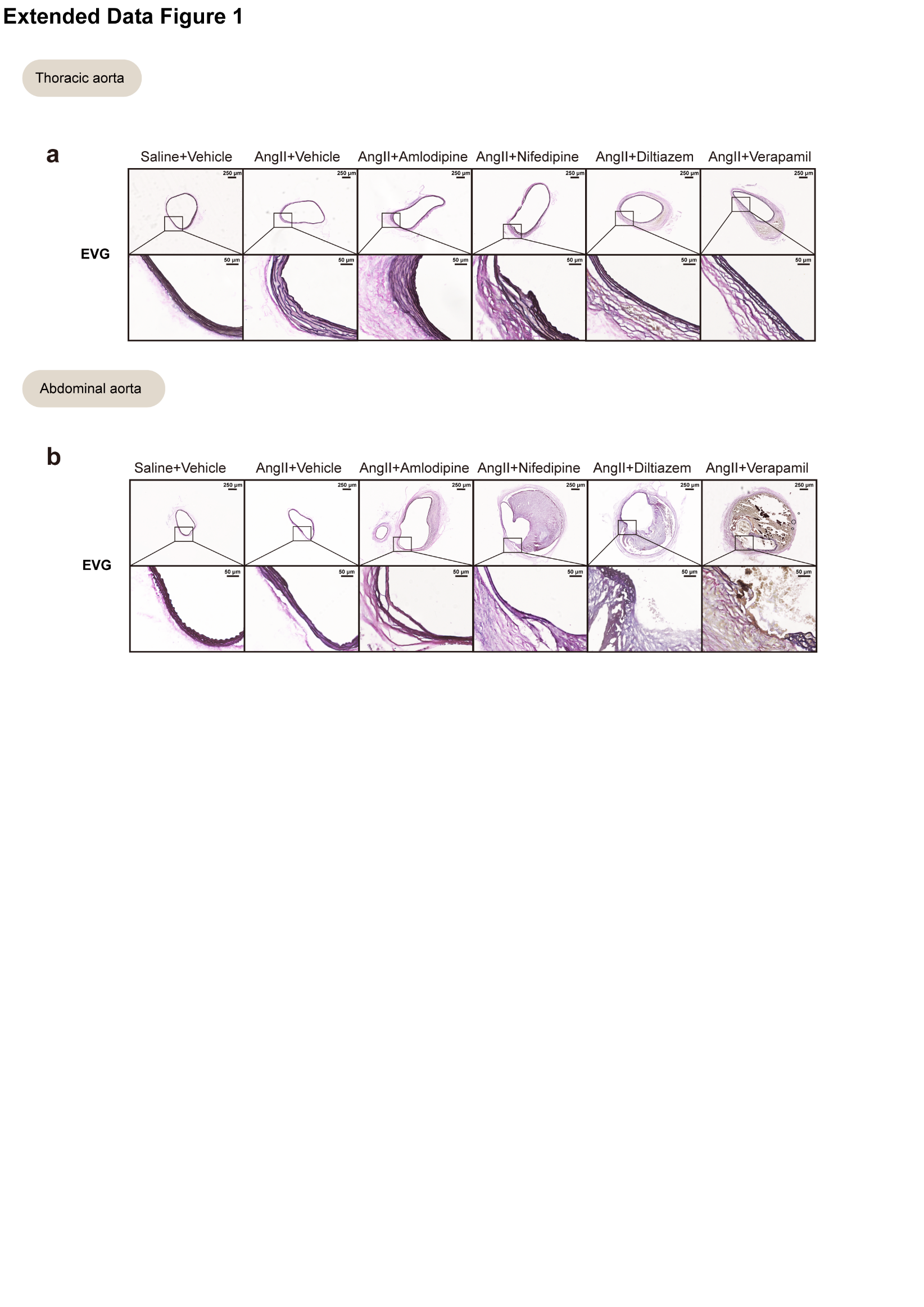
**Extended Data Figure 1 CCBs (amlodipine, nifedipine, diltiazem, verapamil) aggravated thoracic and abdominal aorta elastin degradation in AngII-induced mice model.**

**a-b,** Representative of thoracic and abdominal aorta elastin Van Gieson staining. Scale bar=250/50 μm.

EVG, elastin Van Gieson; Ang II, angiotensin II.

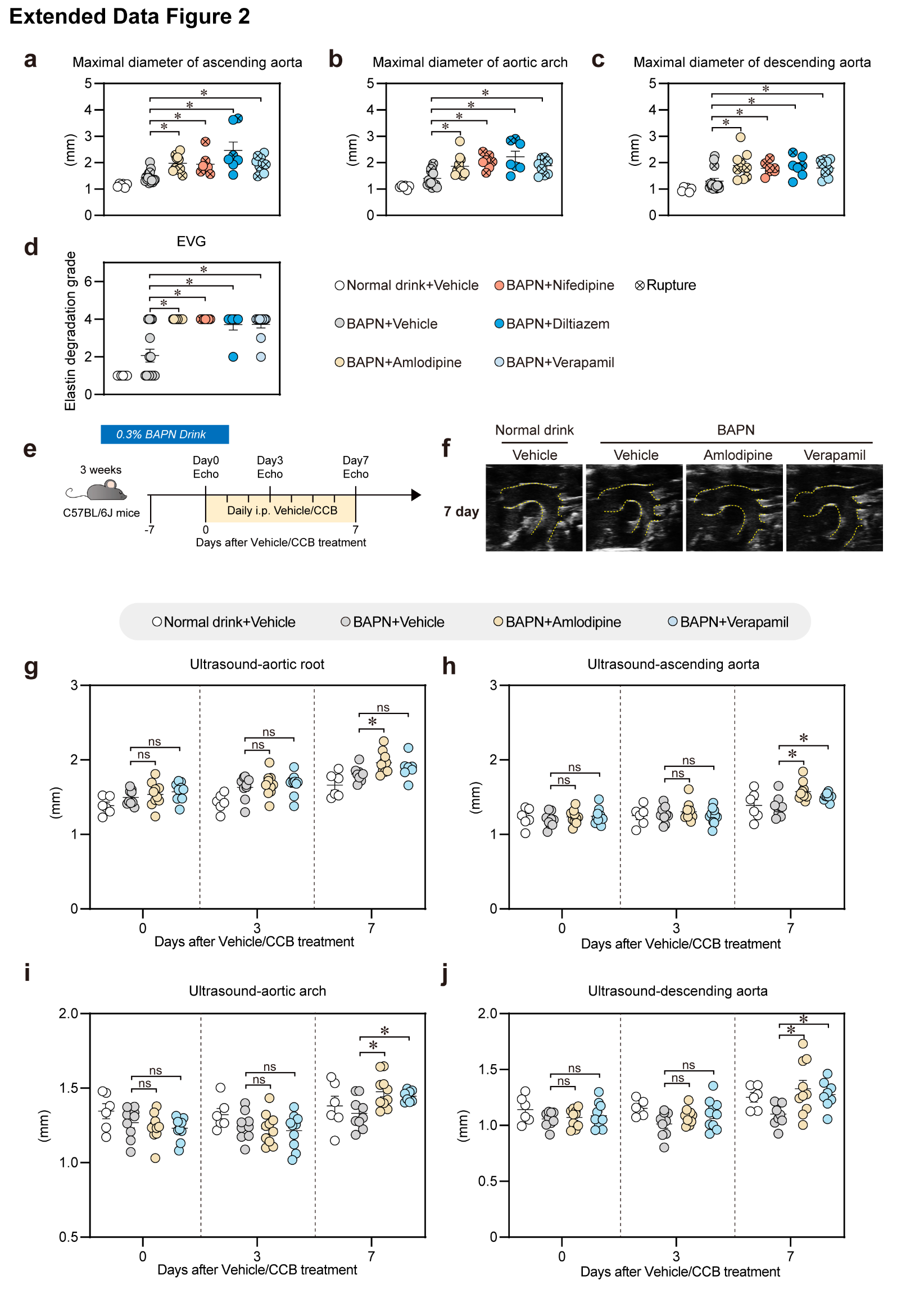

**Extended Data Figure 2 CCBs (amlodipine, nifedipine, diltiazem, verapamil) aggravated BAPN-induced TAAD in mice and aorta stiffness.**

**a-c,** Maximal diameters of ascending aorta, aortic arch and descending aorta.

**d,** Statistical analysis on elastin degradation grade.

**e-j,** Parallel BAPN-model experiment design. (e) 3-week-old mice were treated with 0.3% BAPN in drinking water (0.5 g/kg per day) for 14 days. Normal drink is designed for negative control. After 7 days BAPN drinking, the mice were daily injected with vehicle, amlodipine (1 mg/kg/day) or verapamil (12 mg/kg/day). Groups: Normal drink + vehicle (n=6); BAPN + vehicle (n=10); BAPN + amlodipine (n=10); BAPN + verapamil (n=10). (f) Representative ultrasound images of the thoracic aorta and statistical analysis on (g) aortic root, (h) ascending aorta, (i) aortic arch and (j) descending aorta.

a-d, g-j, Data are the mean ± SEM of each group. **P* < 0.05; ns, not significant. a-c, One-way ANOVA with Dunnett multiple comparisons test. d, Kruskal-Walli’s test comparisons test. g-j, Two-way ANOVA with Dunnett multiple comparisons test.

CCB, calcium channel blocker; BAPN, β-aminopropionitrile.

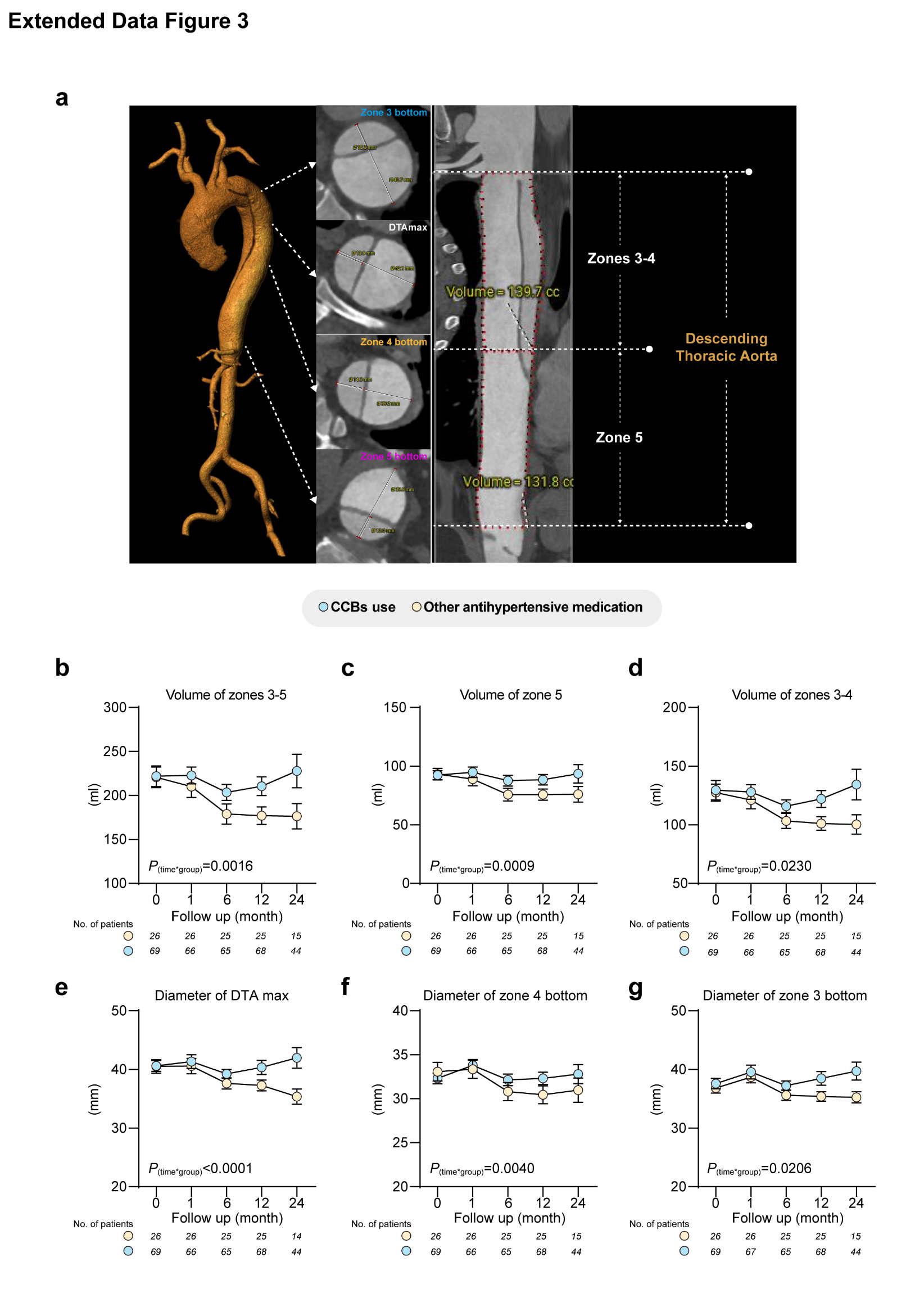

**Extended Data Figure 3 CCBs application limited aortic regression in patients with Type B Aortic Dissection Patients following endovascular therapy.**

**a**, Descending aortic CTA diameters and volumes measurement diagram. Zone 3 (2 cm distal to the left subclavian artery, shown in blue), zone 4 (from the left subclavian artery to the mid - point of the descending thoracic aorta, shown in yellow), and zone 5 (from the mid - point of the DTA to the superior border of the celiac artery, shown in purple). The maximal diameter of the DTA was shown in white.

**b-g**, The volume measurements for (b) zones 3-5, (c) zone 5, and (d) zones 3-4. The diameter measurements for (e) DTA max, (f) zone 4 bottom and (g) zone 3 bottom. The number of patients at each stage is indicated below the corresponding graphs.

b-g, Data are the mean ± SEM of each group. P(time*group) <0.05, statistically significant differences in the trends of diameters and volumes between the two groups over a 2-year follow-up period. b-g, Mixed-effected model was used for the comparisons.

CCB, calcium channel blocker; CTA, computed tomography angiography; DTA max, the max diameter of descending thoracic aorta.

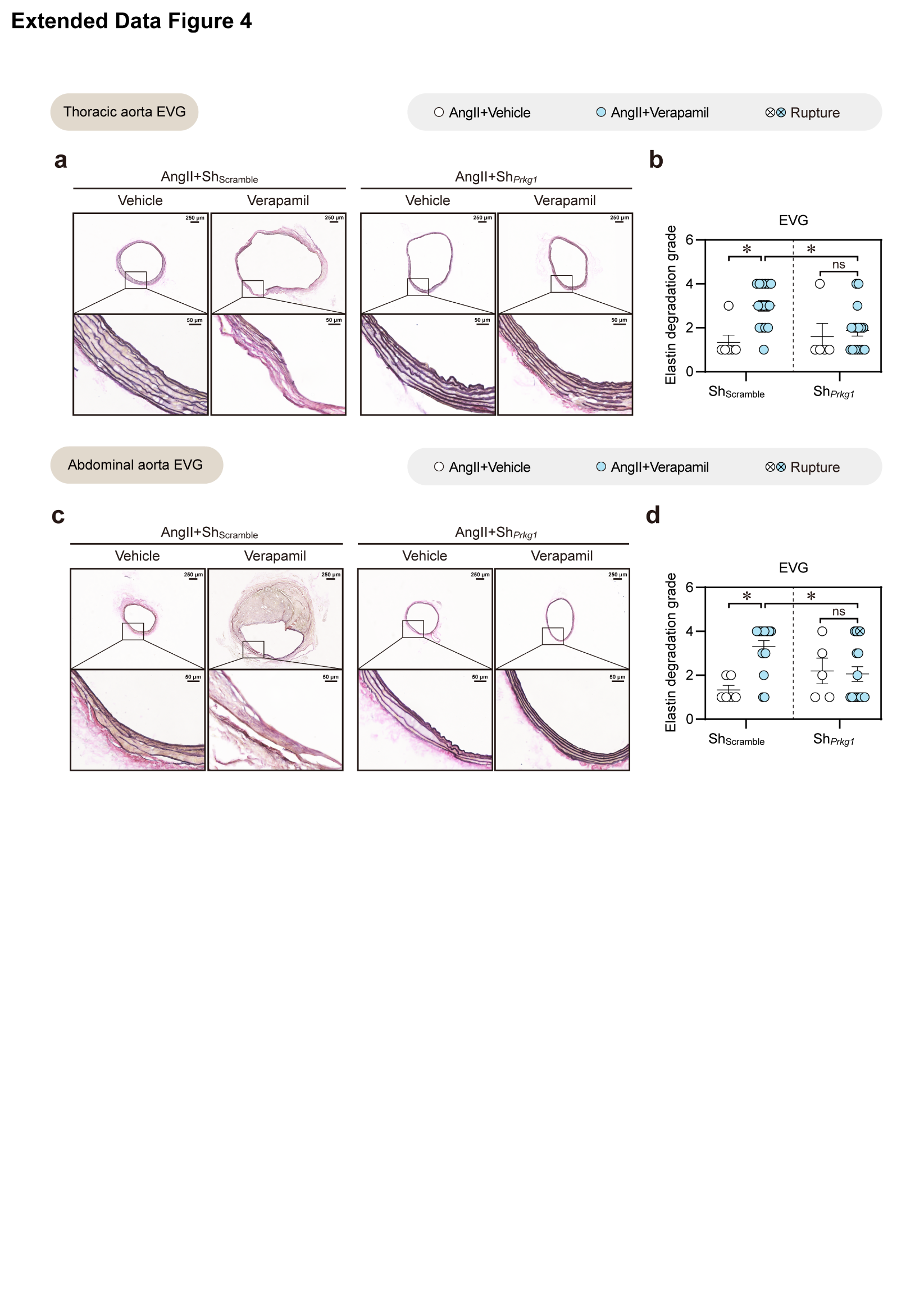

**Extended Data Figure 4** **Silencing of *Prkg1* mitigates CCB-aggravated Elastin degradation in AngII-model.**

**a-d,** Representative of (a) thoracic aorta and (c) abdominal aorta elastin Van Gieson staining. Scale bar=250/50 μm. (b, d) Statistical analysis on elastin degradation grade.

b, d, Data are the mean ± SEM of each group. **P* < 0.05; ns, not significant. b, d, Kruskal-Walli’s test comparisons test.

CCB, calcium channel blocker; Prkg1, protein kinase cGMP-dependent 1; AAV9, adeno-associated virus 9; AngII, angiotensin II.

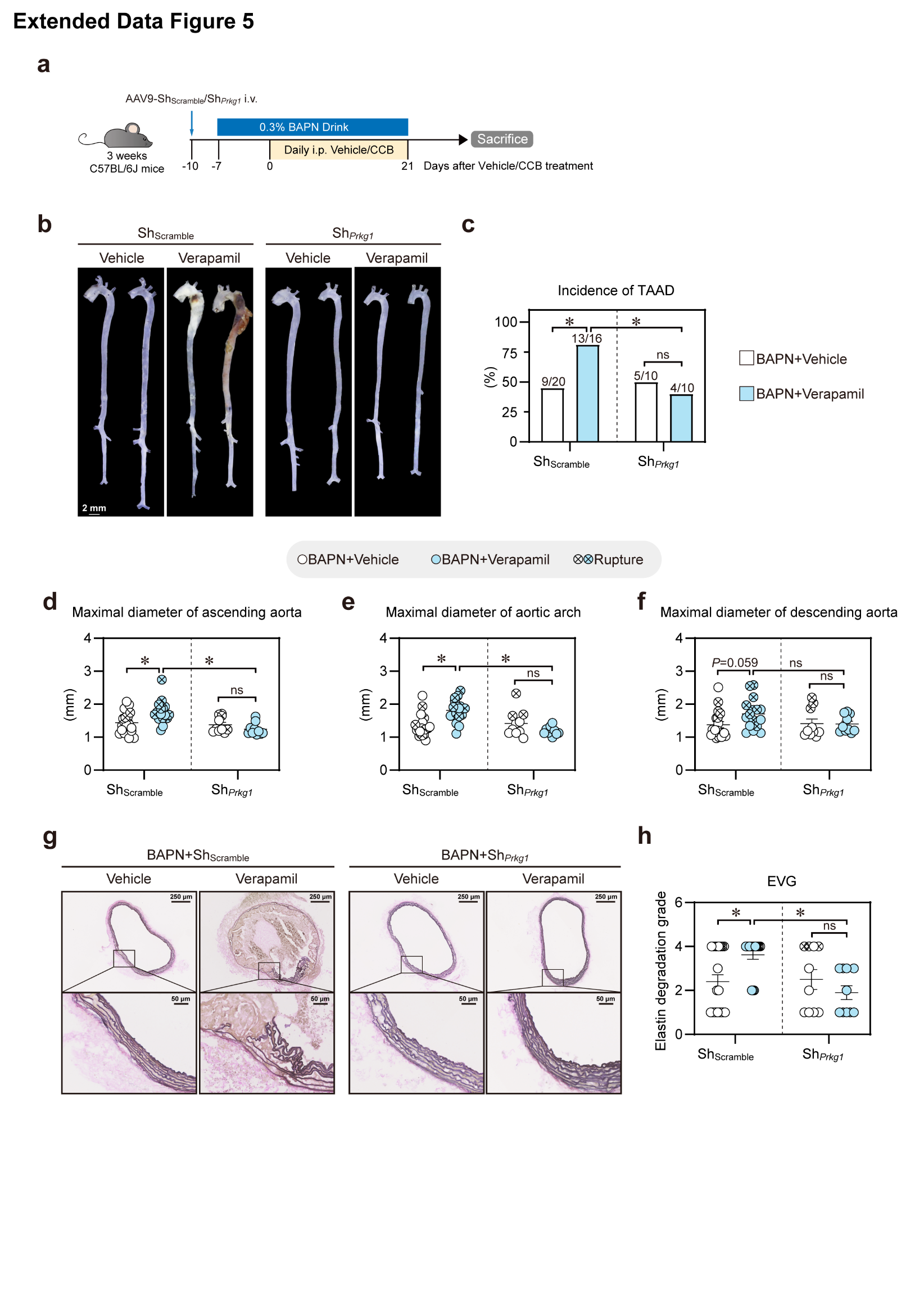
 **Extended Data Figure 5 Silencing of *Prkg1* mitigates CCB-aggravated TAAD in BAPN model.**

**a-c,** 3-week-old C57BL/6J mice were intravenously injected with AAV9-Sh*_Prkg1_* or AAV9-Sh_Scramble_ for 3 days. Then, mice were treated with 0.3% BAPN in drinking water (0.5 g/kg per day) for 28 days. After 7 days of BAPN drinking, the mice were daily injected with vehicle/verapamil (12 mg/kg/day). Groups: Sh_Scramble_ + BAPN + vehicle (n=20); Sh_Scramble_ + BAPN + verapamil (n=16); Sh*_Prkg1_* + BAPN + vehicle (n=10); Sh*_Prkg1_* + BAPN + verapamil (n=10). (b) Representative *ex vivo* morphology of aortas. Scale bar=2 mm. (c) Incidence of TAAD.

**d-f,** Statistical analysis on (d) maximal diameter of ascending aorta, (e) aortic arch and (f) descending aorta.

**g-h,** Representative of (g) thoracic aorta elastin Van Gieson staining. Scale bar=250/50 μm. (h) Statistical analysis on elastin degradation grade.

c, Data are presented as percentage (%). d-f, h, Data are the mean ± SEM of each group. **P* < 0.05; ns, not significant. d-f, Two-way ANOVA with Tukey multiple comparisons test. h, Kruskal-Walli’s test comparisons test.

CCB, calcium channel blocker; Prkg1, protein kinase cGMP-dependent 1; TAAD, thoracic aortic aneurysm and dissection; AAV9, adeno-associated virus 9; BAPN, β-aminopropionitrile.

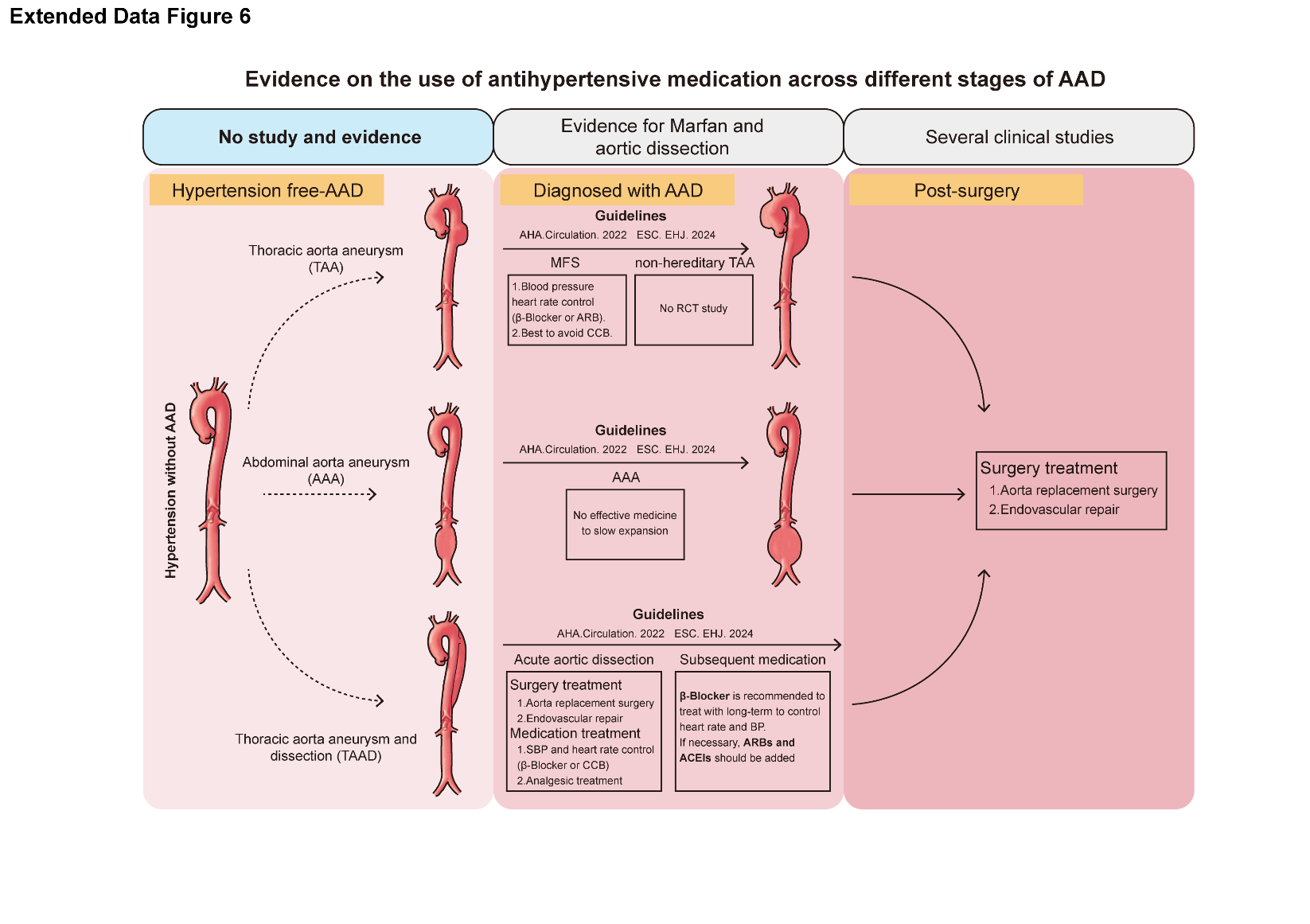
 **Extended Data Figure 6 Evidence on the use of antihypertensive medication across different stage of AAD**

AAD, aortic aneurysm and dissection; MFS, Marfan Syndrome; TAA, thoracic aortic aneurysm; TAAD, thoracic aortic aneurysm and dissection; AAA, abdominal aortic aneurysm; AHA, American Heart Association; ESC, European Society of Cardiology; CCB, calcium channel blockers.

**Extended Data Table 1. Identification of hypertension.**

| Source | Criteria | Data-Field | Coding |
| --- | --- | --- | --- |
| Self-report diagnosed by doctor (high blood pressure) | High blood pressure diagnosed by doctor | 6150 | 4 |
| Self-report during interview | Self-reported high blood pressure during nurse interview | 20002 | 1065, 1072 |
| Hospital data linkage | NA | NA | ICD9:401, 4010, 4011, 4019, 402, 4020, 4021, 4029, 403, 4030, 4031, 4039, 404, 4040, 4041, 4049, 405, 4050, 4051, 4059 |
|  |  |  | ICD10: I10, I11, I110, I119, I12, I120, I129, I13, I130, I131, I132, I139, I15, I150, I151, I152, I158, I159, O10, O100, O101, O102, O103, O104, O109 |

ICD: international classification of diseases.

**Extended Data Table 2. Calcium Channel Blockers medication codes used in the UKB study.**

| Coding | Meaning |
| --- | --- |
| ***Dihydropyridine calcium channel blockers*** | |
| 1140851790 | vasad 5 mg capsule |
| 1140860356 | beta-adalat capsule |
| 1140860358 | tenif capsule |
| 1140860426 | atenolol + nifedipine 50 mg/20 mg m/r capsule |
| 1140861088 | nifedipine |
| 1140861090 | adalat 5 mg capsule |
| 1140861106 | calcilat 10 mg capsule |
| 1140861110 | angiopine 5 mg capsule |
| 1140861114 | nifensar xl 20 mg m/r tablet |
| 1140861120 | coracten sr 10 mg m/r capsule |
| 1140861176 | cardene 20 mg capsule |
| 1140861190 | isradipine |
| 1140861194 | prescal 2.5 mg tablet |
| 1140861276 | lacidipine |
| 1140861282 | motens 2 mg tablet |
| 1140868036 | parmid 10 mg tablet |
| 1140872472 | nimotop 30 mg tablet |
| 1140872568 | nimodipine |
| 1140879802 | amlodipine |
| 1140879810 | nicardipine |
| 1140881702 | adalate 10 mg capsule |
| 1140888646 | felodipine |
| 1140911088 | nifelease 20 mg m/r tablet |
| 1140916930 | calanif 5 mg capsule |
| 1140923572 | adipine mr 10 m/r tablet |
| 1140926188 | unipine xl 30 mg m/r tablet |
| 1140926966 | nimodrel mr 10 m/r tablet |
| 1140927934 | cardilate mr 10 mg m/r tablet |
| 1140927940 | tensipine mr 10 m/r tablet |
| 1140928212 | plendil 2.5 mg m/r tablet |
| 1140928226 | nisoldipine |
| 1140928234 | syscor mr 10 mg m/r tablet |
| 1141145870 | fortipine la 40 m/r tablet |
| 1141150500 | slofedipine 20 mg m/r tablet |
| 1141150538 | nifedotard 20 mr m/r tablet |
| 1141152600 | genalat retard 10 mg m/r tablet |
| 1141153026 | lercanidipine |
| 1141153032 | zanidip 10 mg tablet |
| 1141157140 | nifedipress mr 10 m/r tablet |
| 1141162546 | nivaten retard 10 mg m/r tablet |
| 1141165470 | felodipine + ramipril |
| 1141166752 | coroday mr 20 mg m/r tablet |
| 1141169730 | nifopress retard 20 mg m/r tablet |
| 1141173766 | calchan mr 10 mg m/r tablet |
| 1141187094 | cabren 2.5 mg m/r tablet |
| 1141187962 | kentipine mr 10 mg m/r tablet |
| 1141188152 | felotens xl 5 mg m/r tablet |
| 1141188576 | felogen xl 5 mg m/r tablet |
| 1141188836 | felendil xl 5 mg m/r tablet |
| 1141188920 | keloc sr 5 mg m/r tablet |
| 1141190160 | vascalpha 5 mg m/r tablet |
| 1141190548 | valni 20 retard 20 mg m/r tablet |
| 1141199858 | cardioplen xl 5 mg m/r tablet |
| 1141200400 | amlostin 5 mg tablet |
| 1141200782 | neofel xl 5 mg m/r tablet |
| 1141201814 | parmid xl 5 mg m/r tablet |
| ***Phenylalkylamine calcium channel blockers*** | |
| 1140866460 | half securon sr 120mg m/r tablet |
| 1140866466 | securon 40 mg tablet |
| 1140866484 | geangin 40 mg tablet |
| 1140866546 | berkatens 40 mg tablet |
| 1140866554 | cordilox 40 mg tablet |
| 1140881692 | univer 120 mg m/r capsule |
| 1140888510 | verapamil |
| 1141150926 | verapress mr 240 m/r tablet |
| 1141153316 | tarka 2 mg/180 mg m/r capsule |
| 1141153328 | trandolapril + verapamil hydrochloride |
| 1141169096 | ethimil mr 240 m/r tablet |
| 1141169710 | vertab sr 240 m/r tablet |
| 1141184390 | zolvera 40 mg/5 ml oral solution |
| 1141187056 | ranvera mr 240 mg m/r tablet |
| 1141187774 | vera-til sr 120 mg m/r tablet |
| ***Benzothiazepine calcium channel blockers*** | |
| 1140851730 | calcicard 60 mg tablet |
| 1140861128 | tildiem 60 mg m/r tablet |
| 1140861130 | britiazim 60 mg m/r tablet |
| 1140861136 | angiozem 60 mg m/r tablet |
| 1140861138 | adizem-60 m/r tablet |
| 1140861166 | dilzem sr 60 mg long-acting m/r capsule |
| 1140879806 | diltiazem |
| 1140911698 | slozem 120 mg m/r capsule |
| 1140917428 | angitil sr 90 m/r capsule |
| 1140923618 | kentiazem 60 mg m/r capsule |
| 1140926778 | diltiazem hcl + hydrochlorothiazide 150 mg/12. 5 mg m/r capsule |
| 1140926780 | adizem-xl plus m/r capsule |
| 1141151474 | viazem xl 120 mg m/r capsule |
| 1141153454 | calazem 60 mg m/r tablet |
| 1141156656 | optil 60 mg m/r tablet |
| 1141157136 | dilcardia sr 60 mg m/r capsule |
| 1141167832 | zemtard 120 xl m/r capsule |
| 1141171804 | zildil sr 60 mg m/r capsule |
| 1141174684 | zemret 180 xl m/r capsule |
| 1141175224 | bi-carzem sr 60 mg m/r capsule |
| 1141180238 | horizem sr 90 mg m/r capsule |
| 1141185444 | disogram sr 60 mg m/r capsule |
| ***Other calcium-channel blockers*** | |
| 1141153394 | mibefradil |
| 1141153400 | posicor 50 mg tablet |

**Extended Data Table 3. Other antihypertensive medication excluding CCB code used in the UKB study.**

| **Field ID: 6153; 6177** |  |
| --- | --- |
| coding | meaning |
| ***General Antihypertensive*** | |
| 2 | Self-reported touchscreen question regarding medication for blood pressure |
| **Field ID: 20003** |  |
| coding | meaning |
| ***General Antihypertensive*** | |
| 1140888578 | antihypertensive |
| ***ACE Inhibitor*** | |
| 1140860696 | lisinopril |
| 1140860706 | carace 2.5 mg tablet |
| 1140860714 | zestril 2.5 mg tablet |
| 1140860728 | quinapril |
| 1140860750 | captopril |
| 1140860752 | acepril 12.5 mg tablet |
| 1140860758 | capoten 12.5 mg tablet |
| 1140860776 | innovace 2.5 mg tablet |
| 1140860802 | coversyl 2 mg tablet |
| 1140860806 | ramipril |
| 1140860878 | staril 10 mg tablet |
| 1140860882 | cilazapril |
| 1140860892 | vascace 250 micrograms tablet |
| 1140860904 | trandolapril |
| 1140860912 | gopten 500 micrograms capsule |
| 1140860918 | odrik 500 micrograms capsule |
| 1140888552 | enalapril |
| 1140888556 | fosinopril |
| 1140888560 | perindopril |
| 1140923712 | moexipril |
| 1141151382 | hypapril 12.5 mg tablet |
| 1141164148 | imidapril hydrochloride |
| ***Aldosterone Antagonist*** | |
| 1140866232 | spiroctan-m 200 mg/10ml injection |
| 1140866236 | spironolactone |
| 1140866244 | aldactone 25 mg tablet |
| 1140866262 | rusyde 20 mg tablet |
| 1140866306 | spirospare 25 mg tablet |
| 1140866308 | spiretic 25 mg tablet |
| 1140866312 | spiroctan 25 mg tablet |
| 1140866318 | spirolone 25 mg tablet |
| 1140866396 | aldactide 25 tablet |
| ***Alpha-Beta Blocker*** | |
| 1140879824 | labetalol |
| 1140909368 | carvedilol |
| 1140860610 | hytrin 2 mg tablet |
| ***Alpha-Blocker*** | |
| 1140860690 | cardura 1mg tablet |
| 1140879778 | doxazosin |
| 1140879782 | indoramin |
| 1140879786 | phenoxybenzamine |
| 1140879794 | prazosin |
| 1140879798 | terazosin |
| ***Angiotensin II Receptor Antagonist*** | |
| 1140916356 | losartan |
| 1140916362 | cozaar half strength 25 mg tablet |
| 1141145658 | angiotensin ii receptor antagonist |
| 1141145660 | valsartan |
| 1141145668 | diovan 40 mg capsule |
| 1141152998 | irbesartan |
| 1141153006 | aprovel 75 mg tablet |
| 1141156836 | candesartan cilexetil |
| 1141156846 | amias 2 mg tablet |
| 1141166006 | telmisartan |
| 1141167822 | tensopril 12.5 mg tablet |
| 1141171336 | eprosartan |
| 1141171344 | teveten 300 mg tablet |
| 1141193282 | olmesartan |
| 1141193346 | olmetec 10 mg tablet |
| 1141201040 | co-diovan 80 mg/12.5 mg tablet |
| ***Beta-Blocker*** | |
| 1140860338 | viskaldix tablet |
| 1140860380 | betim 10 mg tablet |
| 1140860382 | blocadren 10 mg tablet |
| 1140860386 | co-betaloc tablet |
| 1140860396 | inderex capsule |
| 1140860398 | kalten capsule |
| 1140860406 | moducren tablet |
| 1140860410 | prestim tablet |
| 1140860434 | monocor 5 mg tablet |
| 1140860492 | emcor 10 mg tablet |
| 1140860498 | celectol 200 mg tablet |
| 1140860564 | dibenyline 10 mg capsule |
| 1140860580 | hypovase 500 mcg tablet |
| 1140860590 | alphavase 500 micrograms tablet |
| 1140860654 | baratol 25 mg tablet |
| 1140860658 | doralese tiltab 20 mg tablet |
| 1140866692 | beta-adrenoceptor blocking drug |
| 1140866704 | angilol 10 mg tablet |
| 1140866712 | cardinol 10 mg tablet |
| 1140866724 | acebutolol |
| 1140866726 | sectral 100 mg capsule |
| 1140866738 | atenolol |
| 1140866756 | tenormin 25 tablet |
| 1140866758 | vasaten 50 mg tablet |
| 1140866764 | apsolol 10 mg tablet |
| 1140866766 | propanix 10 mg tablet |
| 1140866778 | betadur cr 160 mg m/r capsule |
| 1140866782 | beta-prograne 160 mg m/r capsule |
| 1140866784 | berkolol 10 mg tablet |
| 1140866798 | half-betadur cr 80 mg m/r capsule |
| 1140866800 | half-inderal la 80 mg m/r capsule |
| 1140866802 | half beta-prograne 80 mg m/r capsule |
| 1140866804 | inderal 10 mg tablet |
| 1140879760 | bisoprolol |
| 1140879762 | celiprolol |
| 1140879818 | metoprolol |
| 1140879830 | oxprenolol |
| 1140879834 | penbutolol |
| 1140879842 | propranolol |
| 1140879866 | timolol |
| 1140923718 | perdix 7.5 mg tablet |
| 1141156754 | half propatard la 80 mg m/r capsule |
| 1141156808 | propatard la 160 mg m/r capsule |
| 1141164154 | tanatril 5 mg tablet |
| 1141164276 | nebivolol |
| 1141164280 | nebilet 5 mg tablet |
| 1141165476 | triapin mite 2.5 mg/2.5 mg tablet |
| 1141171152 | cardicor 1.25 mg tablet |
| ***Diuretic*** | |
| 1140860334 | trasidrex tablet |
| 1140866072 | hydroflumethiazide |
| 1140866078 | indapamide |
| 1140866090 | methyclothiazide |
| 1140866092 | metolazone |
| 1140866096 | xuret 500 micrograms tablet |
| 1140866102 | polythiazide |
| 1140866104 | nephril 1 mg tablet |
| 1140866122 | bendrofluazide |
| 1140866128 | aprinox 2.5 mg tablet |
| 1140866132 | berkozide 2.5 mg tablet |
| 1140866136 | neo-naclex 5 mg tablet |
| 1140866138 | chlorothiazide |
| 1140866140 | saluric 500 mg tablet |
| 1140866144 | chlorthalidone |
| 1140866146 | hygroton 50 mg tablet |
| 1140866156 | cyclopenthiazide |
| 1140866158 | navidrex 500 mcg tablet |
| 1140866162 | hydrochlorothiazide |
| 1140866164 | esidrex 25 mg tablet |
| 1140866168 | hydrosaluric 25 mg tablet |
| 1140866226 | berkamil 5 mg tablet |
| 1140866352 | navispare tablet |
| 1140866354 | amilmaxco 5/50 tablet |
| 1140866360 | triamaxco tablet |
| 1140866400 | amil-co tablet |
| 1140866402 | dyazide tablet |
| 1140866404 | dytide capsule |
| 1140866410 | kalspare tablet |
| 1140866416 | moduret 25 tablet |
| 1140866420 | moduretic tablet |
| 1140866440 | centyl k m/r tablet |
| 1140866446 | neo-naclex k m/r tablet |
| 1140866450 | bendrofluazide + potassium 2.5 mg/7.7 mmol m/r tablet |
| 1141194794 | bendroflumethiazide |
| 1141194800 | bendroflumethiazide + potassium 2.5 mg/7.7 mmol m/r tablet |
| 1140866340 | delvas tablet |
| 1140866328 | triam-co tablet |
| 1140866324 | triamterene + benzthiazide 50 mg/25 mg capsule |
| 1140866330 | triamterene + chlorthalidone 50 mg/50 mg tablet |
| ***Thyroid Hormone*** | |
| 1140866094 | metenix-5 tablet |
| ***ACE Inhibitor + Diuretic*** | |
| 1140860736 | accuretic tablet |
| 1140860738 | quinalapril + hydrochlorothiazide 10 mg/12.5 mg tablet |
| 1140860764 | captopril + hydrochlorothiazide 25 mg/12.5 mg tablet |
| 1140860784 | innozide tablet |
| 1140860790 | enalapril maleate + hydrochlorothiazide 20 mg/12.5 mg tablet |
| 1140864952 | lisinopril + hydrochlorothiazide 10 mg/12.5 mg tablet |
| 1141180592 | perindopril + indapamide |
| 1141180598 | coversyl plus 4 mg/1.25 mg tablet |
| ***Beta-Blocker + Diuretic*** | |
| 1140860336 | timolol maleate + co-amilozide 10 mg/2.5 mg/25 mg tablet |
| 1140860340 | timolol maleate + bendrofluazide 10 mg/2.5 mg tablet |
| 1140860342 | timolol maleate + bendrofluazide 20 mg/5 mg tablet |
| 1140860348 | atenixco 50 mg/12.5 mg tablet |
| 1140860352 | tenchlor 50 mg/12.5 mg tablet |
| 1140860390 | corgaretic 40 mg tablet |
| 1140860394 | inderetic capsule |
| 1140860402 | lopresoretic tablet |
| 1140860404 | metoprolol tartrate + hydrochlorothiazide 100 mg/12.5 mg tablet |
| 1140860418 | propranolol hydrochloride + bendrofluazide 80 mg/2.5 mg capsule |
| 1140860422 | acebutolol + hydrochlorothiazide 200 mg/12.5 mg tablet |
| 1140864950 | bisoprolol fumarate + hydrochlorothiazide 10 mg/6.25 mg tablet |
| 1141194804 | nadolol + bendroflumethiazide 40 mg/5 mg tablet |
| 1141194808 | timolol maleate + bendroflumethiazide 10 mg/2.5 mg tablet |
| 1141194810 | atenolol + bendroflumethiazide |
| ***Angiotensin II Receptor Antagonist + Diuretic*** | |
| 1141151016 | losartan potassium + hydrochlorothiazide 50 mg/12.5 mg tablet |
| 1141151018 | cozaar-comp 50 mg/12.5 mg tablet |
| 1141172682 | irbesartan + hydrochlorothiazide 150 mg/12.5 mg tablet |
| 1141172686 | coaprovel 150 mg/12.5 mg tablet |
| 1141187788 | telmisartan + hydrochlorothiazide 40 mg/12.5 mg tablet |
| 1141187790 | micardisplus 40 mg/12.5 mg tablet |
| 1141201038 | valsartan + hydrochlorothiazide 80 mg/12.5 mg tablet |
| ***Central Alpha-Agonist + Diuretic*** | |
| 1140860562 | methyldopa + hydrochlorothiazide 250 mg/15 mg tablet |

**Extended Data Table 4. Baseline characteristics of UKB participants according to hypertension status and** **antihypertensive medications use (CCBs or other antihypertensive medications).**

|  | Hypertension | | | | | | No hypertension (n=359,459) | |
| --- | --- | --- | --- | --- | --- | --- | --- | --- |
|  | Without antihypertensive medication (n=43,537) | | With CCBs use (n=34,421) | | With other antihypertensive medication (n=64,461) | |  |  |
| White, % | > 90% | | > 90% | | > 90% | | > 90% | |
| Age, year | 50-70 | | 50-70 | | 50-70 | | 50-70 | |
| BMI, kg/m^2^ | 28.6 (5.00) | | 29.9 (5.28) | | 29.7 (5.29) | | 26.7 (4.37) | |
| Women, % | 22,101 (50.8) | | 14,128 (41.0) | | 32,466 (50.4) | | 204,536 (56.9) | |
| eGFR-MDRD, mL/min/1.73 m² | | 90.8 (30.7) | | 82.9 (31.6) | | 86.6 (31.3) | | 94.8 (31.0) |
| Creatinine, umol/L | | 72.8 (22.7) | | 77.8 (30.7) | | 76.0 (25.5) | | 71.0 (14.2) |
| Urate, umol/L | | 321 (79.8) | | 342 (85.7) | | 345 (84.6) | | 298 (76.1) |
| Albumin, g/L | | 45.3 (2.7) | | 45.4 (2.8) | | 45.0 (2.7) | | 45.2 (2.6) |
| Total cholesterol*^a^*, mmol/L | 5.70 [5.14, 6.51] | | 5.16 [4.36, 5.85] | | 5.29 [4.46, 5.98] | | 5.66 [5.10, 6.43] | |
| Triglycerides*^a^*, mmol/L | 1.53 [1.18, 2.25] | | 1.58 [1.21, 2.28] | | 1.65 [1.25, 2.36] | | 1.48 [1.03, 1.99] | |
| Systolic blood pressure*^a^*, mmHg | 151 [138, 164] | | 148 [137, 160] | | 147 [135, 160] | | 134 [123, 147] | |
| Smoking status, % |  | |  | |  | |  | |
| Never | 22,722 (52.2) | | 16,348 (47.5) | | 33,015 (51.2) | | 204,195 (56.8) | |
| Previous | 15,871 (36.5) | | 14,696 (42.7) | | 25,976 (40.3) | | 116,184 (32.3) | |
| Current | 4,944 (11.4) | | 3,377 (9.8) | | 5,470 (8.5) | | 39,080 (10.9) | |
| Alcohol drinker status, % |  | |  | |  | |  | |
| Never | 1,724 (4.0) | | 1,975 (5.7) | | 3,575 (5.5) | | 15,087 (4.2) | |
| Previous | 1,802 (4.1) | | 1,679 (4.9) | | 2,921 (4.5) | | 11,658 (3.2) | |
| Current | 40,011 (91.9) | | 30,767 (89.4) | | 57,965 (89.9) | | 332,714 (92.6) | |
| Education level, % |  | |  | |  | |  | |
| Higher education | ~30% | | ~30% | | ~30% | | ~30% | |

*^a^* Data are medians (interquartile range, [Q1, Q3]).

CCB, calcium channel blocker; BMI, body mass index.

**Extended Data Table 5. ICD codes used for identification of disease outcomes.**

| Disease outcome | Definition |
| --- | --- |
| AAD | Self-reported non-cancer illness code (Data-Field:20002): 1492 [aortic aneurysm], 1592 [aortic dissection] |
|  | ICD10: I710, I711, I712, I713, I714, I715, I716, I718, I719 |
| TAAD | ICD10: I710, I711, I712 |
| AAA | ICD10: I713, I714 |

ICD, international classification of diseases; AAD, aortic aneurysm and dissection; TAAD, thoracic aortic aneurysm and dissection; AAA, abdominal aortic aneurysm.

**Extended Data Table 6. Association of CCBs and AAD risk among UKB participants with hypertension and other antihypertensive medications (n=98,882).**

|  | No. cases/person years | Hazard ratio (95% CI) *^a^* | *P* value |
| --- | --- | --- | --- |
| Hypertension with other antihypertensive medication (Ref.) | 1,002 / 819,548 | Ref. |  |
| Hypertension with CCBs use | 756 / 429,687 | 1.23 (1.09, 1.38) | <0.001 |

*^a^* Cox model was adjusted for age (years), sex (men; women), BMI (kg/m^2^), total cholesterol (mmol/L), triglycerides (mmol/L), smoking status (never; previous; current), education (College or University degree; A levels or equivalent; O levels or equivalent; None of the above), ethnicity (white; or not), and systolic blood pressure (mmHg).

AAD, aortic aneurysm and dissection; CCB, calcium channel blocker; BMI, body mass index.

**Extended Data Table 7. Association of CCBs and all-cause mortality among UKB participants with prevalent AAD (n=509).**

|  | No. cases/person years | Hazard ratio (95% CI) *^a^* | *P* value |
| --- | --- | --- | --- |
| Hypertension without antihypertensive medication (ref) | 46 / 1,593 | Ref. |  |
| CCBs use | 85 / 1,864 | 1.52 (1.04, 2.21) | 0.03 |
| Hypertension with other antihypertensive medication | 83 / 2,260 | 1.28 (0.89, 1.85) | 0.18 |

*^a^* Cox model was adjusted for age (years), sex (men; women), BMI (kg/m^2^), total cholesterol (mmol/L), triglycerides (mmol/L), smoking status (never; previous; current), education (College or University degree; A levels or equivalent; O levels or equivalent; None of the above), ethnicity (white; or not), and systolic blood pressure (mmHg).

AAD, aortic aneurysm and dissection; CCB, calcium channel blocker; BMI, body mass index.

**Extended Data Table 8. Sensitivity analysis - association of hypertension status and antihypertensive medication use (CCBs or other antihypertensive medication excluding CCB use) with AAD/AAD subtypes risks among UKB participants (n=501,878) *^a^*.**

| AAD | No. Cases/person years | HR (95% CI) *^a^* | P value |
| --- | --- | --- | --- |
| Hypertension without antihypertensive medication (ref.) | 566 / 649,485 | 1 (ref.) |  |
| Hypertension with CCBs use | 799 / 453,312 | 1.32 (1.18, 1.47) | <0.001 |
| Hypertension with other antihypertensive medication | 1,119 / 881,440 | 1.09 (0.98, 1.21) | 0.11 |
| No hypertension | 1,843 / 4,552,599 | 0.65 (0.59, 0.71) | <0.001 |
| **TAAD** | No. Cases/person years | HR (95% CI) *^a^* | P value |
| Hypertension without antihypertensive medication (ref.) | 202 / 651,012 | 1 (ref.) |  |
| Hypertension with CCBs use | 228 / 455,756 | 1.26 (1.04, 1.53) | 0.02 |
| Hypertension with other antihypertensive medication | 378 / 884,555 | 1.19 (0.98, 1.41) | 0.07 |
| No hypertension | 712 / 4,557,043 | 0.59 (0.50, 0.69) | <0.001 |
| **AAA** | No. Cases/person years | HR (95% CI) *^a^* | P value |
| Hypertension without antihypertensive medication (ref.) | 343 / 650,315 | 1 (ref.) |  |
| Hypertension with CCBs use | 530 / 454,319 | 1.33 (1.16, 1.53) | <0.001 |
| Hypertension with other antihypertensive medication | 691 / 883,077 | 1.08 (0.94, 1.23) | 0.26 |
| No hypertension | 1,016 / 4,555,548 | 0.59 (0.52, 0.67) | <0.001 |

*^a^* Hypertension was applied with a broader definition through self-reported hypertension during nurse interview, self-reported questionnaires regarding high blood pressure diagnosed by doctor, hospital data via ICD codes, and self-reported antihypertensive medications during interview or questionnaires in the sensitivity analysis. Cox model was adjusted for age (years), sex (men; women), BMI (kg/m^2^), total cholesterol (mmol/L), triglycerides (mmol/L), smoking status (never; previous; current), education (College or University degree; A levels or equivalent; O levels or equivalent; None of the above), ethnicity (white; or others), and systolic blood pressure (mmHg).

AAD, aortic aneurysm and dissection; TAAD, thoracic aortic aneurysm and dissection; AAA, abdominal aortic aneurysm; CCB, calcium channel blocker; HR, hazard ratio; BMI, body mass index; ICD, international classification of diseases.

**Extended Data Table 9. Sensitivity analysis - association of hypertension status and antihypertensive medication use (CCBs or other antihypertensive medication excluding CCB use) with AAD/AAD subtypes risks among UKB participants using Fine & gray Models for competing risk of death (n=501,878) *^a^*.**

| AAD | No. Cases/person years | HR (95% CI) *^a^* | P value |
| --- | --- | --- | --- |
| Hypertension without antihypertensive medication (ref.) | 465 / 565,794 | 1 (ref.) |  |
| Hypertension with CCBs use | 756 / 429,687 | 1.26 (1.12, 1.42) | <0.001 |
| Hypertension with other antihypertensive medication | 1,002 / 819,548 | 1.04 (0.93, 1.16) | 0.54 |
| No hypertension | 2,104 / 472,1807 | 0.66 (0.59, 0.73) | <0.001 |
| **TAAD** | No. Cases/person years | HR (95% CI) *^a^* | P value |
| Hypertension without antihypertensive medication (ref.) | 177 / 567,019 | 1 (ref.) |  |
| Hypertension with CCBs use | 222 / 431,992 | 1.21 (1.01, 1.48) | 0.05 |
| Hypertension with other antihypertensive medication | 334 / 822,396 | 1.07 (0.88, 1.29) | 0.50 |
| No hypertension | 787 / 4,721,807 | 0.60 (0.51, 0.72) | <0.001 |
| **AAA** | No. Cases/person years | HR (95% CI) *^a^* | P value |
| Hypertension without antihypertensive medication (ref.) | 269 / 566,521 | 1 (ref.) |  |
| Hypertension with CCBs use | 494 / 430,655 | 1.25 (1.07, 1.45) | 0.005 |
| Hypertension with other antihypertensive medication | 617 / 821,018 | 1.00 (0.86, 1.16) | 0.37 |
| No hypertension | 1,200 / 4,725,064 | 0.70 (0.61, 0.80) | <0.001 |

*^a^* Fine & gray model was adjusted for age (years), sex (men; women), BMI (kg/m^2^), total cholesterol (mmol/L), triglycerides (mmol/L), smoking status (never; previous; current), education (College or University degree; A levels or equivalent; O levels or equivalent; None of the above), ethnicity (white; or others), and systolic blood pressure (mmHg).

AAD, aortic aneurysm and dissection; TAAD, thoracic aortic aneurysm and dissection; AAA, abdominal aortic aneurysm; CCB, calcium channel blocker; HR, hazard ratio; BMI, body mass index.

**Extended Data Table 10. Sensitivity analysis - association of hypertension status and antihypertensive medication use (CCBs or other antihypertensive medication excluding CCB use) with AAD/AAD subtypes risks among UKB participants after removing prevalent coronary artery disease (n=469,927) *^a^*.**

| AAD | No. Cases/person years | HR (95% CI) *^a^* | P value |
| --- | --- | --- | --- |
| Hypertension without antihypertensive medication (ref.) | 358 / 526,674 | 1 (ref.) |  |
| Hypertension with CCBs use | 502 / 370,384 | 1.25 (1.09, 1.44) | 0.002 |
| Hypertension with other antihypertensive medication | 691 / 712,948 | 1.08 (0.95, 1.23) | 0.23 |
| No hypertension | 1,681 / 4,531,435 | 0.65 (0.58, 0.73) | <0.001 |
| **TAAD** | No. Cases/person years | HR (95% CI) *^a^* | P value |
| Hypertension without antihypertensive medication (ref.) | 141 / 527,750 | 1 (ref.) |  |
| Hypertension with CCBs use | 168 / 371,741 | 1.26 (1.03, 1.59) | 0.04 |
| Hypertension with other antihypertensive medication | 249 / 714,811 | 1.09 (0.89, 1.35) | 0.40 |
| No hypertension | 787 / 4,535,513 | 0.59 (0.49, 0.72) | <0.001 |
| **AAA** | No. Cases/person years | HR (95% CI) *^a^* | P value |
| Hypertension without antihypertensive medication (ref.) | 204 / 527,203 | 1 (ref.) |  |
| Hypertension with CCBs use | 311 / 371,101 | 1.22 (1.01, 1.46) | 0.03 |
| Hypertension with other antihypertensive medication | 402 / 714,024 | 1.04 (0.87, 1.23) | 0.66 |
| No hypertension | 948 / 4,534,063 | 0.68 (0.59, 0.80) | <0.001 |

*^a^* Participants with prevalent coronary artery diseases in this sensitivity analysis were further excluded. Cox model was adjusted for age (years), sex (men; women), BMI (kg/m^2^), total cholesterol (mmol/L), triglycerides (mmol/L), smoking status (never; previous; current), education (College or University degree; A levels or equivalent; O levels or equivalent; None of the above), ethnicity (white; or others), and systolic blood pressure (mmHg).

AAD, aortic aneurysm and dissection; TAAD, thoracic aortic aneurysm and dissection; AAA, abdominal aortic aneurysm; CCB, calcium channel blocker; HR, hazard ratio; BMI, body mass index.

**Extended Data Table 11. Sensitivity analysis - association of anti-hypertensive medication use with AAD Among UKB participants further adjusted for eGFR (n=501,878) *^a^*.**

| AAD | No. Cases/person years | HR (95% CI) *^a^* | P value |
| --- | --- | --- | --- |
| Hypertension without antihypertensive medication (ref.) | 465 / 565,794 | 1 (Ref.) |  |
| Hypertension with exclusive CCBs use | 756 / 429,687 | 1.31 (1.16, 1.48) | <0.001 |
| Hypertension with other antihypertensive medication | 1,002 / 819,548 | 1.08 (0.96, 1.20) | 0.20 |
| No hypertension | 2,104 / 4,721,807 | 0.64 (0.58, 0.71) | <0.001 |
| **TAAD** | No. Cases/person years | HR (95% CI) *^a^* | P value |
| Hypertension without antihypertensive medication (ref.) | 177 / 567,019 | 1 (Ref.) |  |
| Hypertension with exclusive CCBs use | 222 / 431,992 | 1.27 (1.00, 1.61) | 0.05 |
| Hypertension with other antihypertensive medication | 334 / 822,396 | 1.15 (0.92, 1.43) | 0.22 |
| No hypertension | 787 / 4,721,807 | 0.64 (0.52, 0.78) | <0.001 |
| **AAA** | No. Cases/person years | HR (95% CI) *^a^* | P value |
| Hypertension without antihypertensive medication (ref.) | 269 / 566,521 | 1 (Ref.) |  |
| Hypertension with exclusive CCBs use | 494 / 430,655 | 1.32 (1.13, 1.53) | <0.001 |
| Hypertension with other antihypertensive medication | 617 / 821,018 | 1.06 (0.92, 1.23) | 0.44 |
| No hypertension | 1,200 / 4,725,064 | 0.66 (0.58, 0.76) | <0.001 |

*^a^* Cox model was adjusted for age (years), sex (men; women), BMI (kg/m2), total cholesterol (mmol/L), triglycerides (mmol/L), smoking status (never; previous; current), education (College or University degree; A levels or equivalent; O levels or equivalent; None of the above), ethnicity (white; or others), systolic blood pressure (mmHg) and eGFR.

AAD, aortic aneurysm and dissection; TAAD, thoracic aortic aneurysm and dissection; AAA, abdominal aortic aneurysm; CCB, calcium channel blocker; HR, hazard ratio; BMI, body mass index.

**Extended Data Table 12. Sensitivity analysis - association of hypertension status and antihypertensive medication use with AAD/AAD subtypes risks among UKB participants after removing participants with hypertension in combination of both concurrent use of CCBs and other antihypertensive medication (n=496,256) *^a^*.**

| AAD | No. Cases/person years | HR (95% CI) *^a^* | P value |
| --- | --- | --- | --- |
| Hypertension without antihypertensive medication (ref.) | 465 / 565,794 | 1 (ref.) |  |
| Hypertension with exclusive CCBs use | 644 / 359,676 | 1.33 (1.18, 1.51) | <0.001 |
| Hypertension with other antihypertensive medication | 1,002 / 819,548 | 1.08 (0.97, 1.21) | 0.16 |
| No hypertension | 2,104 / 4,721,807 | 0.64 (0.58, 0.71) | <0.001 |
| **TAAD** | No. Cases/person years | HR (95% CI) *^a^* | P value |
| Hypertension without antihypertensive medication (ref.) | 177 / 567,019 | 1 (ref.) |  |
| Hypertension with exclusive CCBs use | 178 / 361,671 | 1.18 (0.95, 1.46) | 0.13 |
| Hypertension with other antihypertensive medication | 334 / 822,396 | 1.09 (0.90, 1.31) | 0.37 |
| No hypertension | 787 / 4,726,960 | 0.60 (0.50, 0.71) | <0.001 |
| **AAA** | No. Cases/person years | HR (95% CI) *^a^* | P value |
| Hypertension without antihypertensive medication (ref.) | 269 / 566,521 | 1 (ref.) |  |
| Hypertension with exclusive CCBs use | 434 / 360,447 | 1.37 (1.18, 1.60) | <0.001 |
| Hypertension with other antihypertensive medication | 617 / 821,018 | 1.06 (0.92, 1.23) | 0.40 |
| No hypertension | 1,200 / 4,725,064 | 0.66 (0.58, 0.76) | <0.001 |

*^a^* Participants with hypertension in combination of both concurrent use of CCBs and other antihypertensive medication in this sensitivity analysis were further excluded. Cox model was adjusted for age (years), sex (men; women), BMI (kg/m^2^), total cholesterol (mmol/L), triglycerides (mmol/L), smoking status (never; previous; current), education (College or University degree; A levels or equivalent; O levels or equivalent; None of the above), ethnicity (white; or others), and systolic blood pressure (mmHg).

AAD, aortic aneurysm and dissection; TAAD, thoracic aortic aneurysm and dissection; AAA, abdominal aortic aneurysm; CCB, calcium channel blocker; HR, hazard ratio; BMI, body mass index.

**Extended Data Table 13. Characteristics of mice (AngII-model) infused with vehicle, amlodipine, nifedipine, diltiazem and verapamil for 21 days.**

| Groups (♂) | Saline | AngII | | | | |
| --- | --- | --- | --- | --- | --- | --- |
|  | Vehicle | Vehicle | Amlodipine | Nifedipine | Diltiazem | Verapamil |
| No. | N=7 | N=13 | N=12 | N=12 | N=11 | N=11 |
| Weight -7 days (g) | 29.72 ± 0.39 | 28.35 ± 0.61 | 28.17 ± 0.68 | 29.50 ± 0.25 | 29.61 ± 0.23 | 29.01 ± 0.93 |
| Weight 0 day (g) | 29.45 ± 0.79 | 28.38 ± 0.65 | 27.99 ± 0.62 | 30.17 ± 0.28 | 29.08 ± 0.53 | 28.40 ± 0.78 |
| Weight 7 days (g) | 30.52 ± 0.82 | 29.08 ± 0.69 | 28.55 ± 0.77 | 29.24 ± 0.38 | 29.23 ± 0.45 | 29.77 ± 1.10 |
| Weight 14 days (g) | 30.26 ± 0.70 | 29.95 ± 0.67 | 29.24 ± 0.75 | 29.60 ± 0.40 | 29.32 ± 0.42 | 31.08 ± 1.09 |
| Weight 21 days (g) | 30.81 ± 0.54 | 29.96 ± 0.49 | 29.68 ± 0.59 | 30.15 ± 0.31 | 30.13 ± 0.27 | 31.20 ± 0.61 |
| TC (mg/dl) | 177.28 ±  5.99 | 185.43 ±  16.94 | 184.25 ±  12.79^ns^ | 173.92 ±  10.77^ns^ | 170.02 ±  7.68^ns^ | 175.28 ±  9.78^ns^ |
| TG (mg/dl) | 87.22 ±  5.70 | 93.56 ±  8.89 | 91.97 ±  9.56^ns^ | 89.01 ±  6.40^ns^ | 88.09 ±  5.43^ns^ | 91.29 ± 15.46^ns^ |

Each group weight was compared using Kruskal-Wallis test with Dunn's multiple comparisons test.

Comparison of AngII + CCB group *vs.* AngII + Vehicle group.

For TC and TG, Kruskal-Wallis test with Dunn's multiple comparisons test.

Data are presented as the mean ± SEM; ns, not significant.

TC, total cholesterol; TG, triglyceride.

**Extended Data Table 14. Characteristics of mice (Elastase-AAA model) infused with control, vehicle, amlodipine, nifedipine, diltiazem and verapamil for 14 days.**

| Groups (♂) | PBS | Elastase | | | | |
| --- | --- | --- | --- | --- | --- | --- |
|  | Vehicle | Vehicle | Amlodipine | Nifedipine | Diltiazem | Verapamil |
| No. | N=3 | N=9 | N=11 | N=10 | N=12 | N=9 |
| Weight 0 day (g) | 19.81 ± 0.31 | 19.86 ± 0.22 | 20.44 ± 0.32 | 19.84 ± 0.28 | 20.84 ± 0.20 | 19.78 ± 0.28 |
| Weight 7 days (g) | 20.77 ± 0.35 | 21.38 ± 0.28 | 21.47 ± 0.34 | 21.82 ± 0.42 | 21.62 ± 0.32 | 21.43 ± 0.35 |
| Weight 14 days (g) | 22.11 ± 0.45 | 22.66 ± 0.30 | 22.72 ± 0.30 | 22.67 ± 0.42 | 22.82 ± 0.25 | 22.61 ± 0.32 |

Each group weight was compared using Kruskal-Wallis test with Dunn's multiple comparisons test.

Data are presented as the mean ± SEM.

**Extended Data Table 15. Characteristics of mice (BAPN-TAAD model) infused with vehicle, amlodipine, nifedipine, diltiazem and verapamil for 21 days.**

| Groups (♂) | Water | BAPN | | | | |
| --- | --- | --- | --- | --- | --- | --- |
|  | Vehicle | Vehicle | Amlodipine | Nifedipine | Diltiazem | Verapamil |
| No. | N=6 | N=15 | N=10 | N=7 | N=7 | N=10 |
| Weight -7 days (g) | 11.69 ± 0.27 | 12.48 ± 0.27 | 12.15 ± 0.15 | 12.16 ± 0.25 | 11.43 ± 0.47 | 11.80 ± 0.25 |
| Weight 0 day (g) | 18.01 ± 0.16 | 16.24 ± 0.43 | 16.18 ± 0.22 | 16.59 ± 0.53 | 16.34 ± 0.50 | 15.44 ± 0.26 |
| Weight 7 days (g) | 19.62 ± 0.40 | 17.62 ± 0.41 | 18.00 ± 0.32 | 17.05 ± 0.52 | 17.64 ± 0.37 | 17.31 ± 0.32 |
| Weight 14 days (g) | 21.23 ± 0.33 | 18.54 ± 0.51 | 19.12 ± 0.52 | 19.85 ± 0.41 | 19.27 ± 0.47 | 18.25 ± 0.14 |
| Weight 21 days (g) | 22.46 ± 0.40 | 19.6 ± 0.40 | 20.63 ± 0.68 | 20.85 ± 0.44 | 20.30 ± 0.52 | 19.45 ± 0.72 |
| Water -7 days (ml/g/d) | 0.257 | 0.187 | 0.197 | 0.188 | 0.188 | 0.229 |
| Water 0 day (ml/g/d) | 0.259 | 0.181 | 0.204 | 0.172 | 0.188 | 0.207 |
| Water 7 days (ml/g/d) | 0.255 | 0.189 | 0.194 | 0.176 | 0.211 | 0.217 |
| Water 14 days (ml/g/d) | 0.224 | 0.200 | 0.209 | 0.171 | 0.185 | 0.196 |
| Water 21 days (ml/g/d) | 0.230 | 0.192 | 0.204 | 0.192 | 0.197 | 0.206 |

Each group weight was compared using Kruskal-Wallis test with Dunn's multiple comparisons test.

Weight data are presented as the mean ± SEM.

Water intake data were presented for per unit of body weight and per day.

**Extended Data Table 16. The information of excluded 25 patients.**

| Number | Gender | Age | Hypertension | Event time  (post TEVAR) | Reason for exclusion |
| --- | --- | --- | --- | --- | --- |
| Patient 1 | / | 50-60 | / | / | Death |
| Patient 2 | / | 60-70 | / | / | Death |
| Patient 3 | / | 60-70 | / | / | Death |
| Patient 4 | / | 50-60 | / | / | Death |
| Patient 5 | / | 70-80 | / | / | Death |
| Patient 6 | / | 50-60 | / | / | Brain disease |
| Patient 7 | / | 70-80 | / | / | Brain disease |
| Patient 8 | / | 70-80 | / | / | Brain and Respiratory disease |
| Patient 9 | / | 70-80 | / | / | Respiratory disease |
| Patient 10 | / | 40-50 | / | / | Complication |
| Patient 11 | / | 40-50 | / | / | Complication |
| Patient 12 | / | 40-50 | / | / | Complication |
| Patient 13 | / | 70-80 | / | / | Complication |
| Patient 14 | / | 40-50 | / | / | Complication |
| Patient 15 | / | 40-50 | / | / | Complication |
| Patient 16 | / | 40-50 | / | / | Complication |
| Patient 17 | / | 70-80 | / | / | Complication |
| Patient 18 | / | 60-70 | / | / | Other reason |
| Patient 19 | / | 60-70 | / | / | / |
| Patient 20 | / | 60-70 | / | / | / |
| Patient 21 | / | 60-70 | / | / | / |
| Patient 22 | / | 60-70 | / | / | / |
| Patient 23 | / | 50-60 | / | / | / |
| Patient 24 | / | 60-70 | / | / | / |
| Patient 25 | / | 50-60 | / | / | / |

**Extended Data Table 17. Patients’ baseline characteristics and blood, urine examination.**

| Number of patients | | CCBs use  (n=69) | Other antihypertensive medication  (n=26) | *P* value |
| --- | --- | --- | --- | --- |
| Demographic characteristics | |  |  |  |
|  | Age (years) ^a^ | 50-60 | 50-60 | 0.652 |
|  | Male, n (%) | 59 (85.5%) | 18 (69.2%) | 0.084 |
|  | BMI (kg/m^2^) ^a^ | 25.91 ± 0.38 | 27.11 ± 0.84 | 0.150 |
| Clinical history | |  |  |  |
|  | Hypertension, n (%) | 69 (100%) | 26 (100%) | 1.000 |
|  | SBP (mmHg) ^a^ | 128.84 ± 2.10 | 132.08 ± 2.79 | 0.400 |
|  | Family genetic history, n (%) | 3 (4.3%) | 1 (3.8%) | 1.000 |
|  | Diabetes, n (%) | 3 (4.3%) | 1 (3.8%) | 1.000 |
|  | Hyperlipidemia, n (%) | 3 (4.3%) | 0 | 0.559 |
|  | Coronary heart disease, n (%) | 3 (4.3%) | 0 | 0.559 |
|  | MI | 0 | 0 |  |
|  | TIA | 0 | 0 |  |
|  | Cerebral infarction | 0 | 0 |  |
| Blood routine ^a^ | |  |  |  |
|  | RBC (×10^12^/L) | 4.47 ± 0.07 | 4.37 ± 0.12 | 0.424 |
|  | WBC (×10^9^/L) | 9.36 ± 0.36 | 9.83 ± 0.50 | 0.486 |
|  | PLT (×10^9^/L) | 216.35 ± 10.22 | 232.85 ± 18.34 | 0.414 |
|  | HB (g/L) | 136.68 ± 1.88 | 133.37 ± 2.98 | 0.356 |
| Urine routine ^a^ | |  |  |  |
|  | U-RBC (/hpf) | 37.48 ± 14.23 | 31.65 ± 8.72 | 0.731 |
|  | U-WBC (/hp) | 35.95 ± 16.74 | 11.44 ± 3.97 | 0.384 |
|  | U-PRO (mg/L) | 1.24 ± 0.05 | 1.40 ± 0.10 | 0.119 |
|  | U-PH | 6.12 ± 0.09 | 6.12 ± 0.13 | 0.989 |
| Blood Biochemistry ^a^ | |  |  |  |
|  | Creatinine (μmol/L) | 76.51 ± 2.41 | 67.67 ± 3.26 | 0.048 |
|  | ALT/GPT (U/L) | 29.15 ± 2.60 | 27.48 ± 4.35 | 0.946 |
|  | AST/GOT (U/L) | 23.82 ± 1.18 | 24.55 ± 2.64 | 0.615 |
|  | TBIL (μmol/L) | 16.21 ± 1.11 | 17.58 ± 1.36 | 0.785 |
|  | DBIL (μmol/L) | 4.14 ± 0.33 | 5.00 ± 0.45 | 0.342 |
|  | IBIL (μmol/L) | 12.13 ± 0.92 | 12.33 ± 1.19 | 0.894 |
| Coagulation examination ^a^ | |  |  |  |
|  | APTT (s) | 30.74 ± 0.58 | 30.53 ± 1.34 | 0.890 |
|  | PT (s) | 11.94 ± 0.15 | 11.98 ± 0.29 | 0.904 |
|  | TT (s) | 15.58 ± 0.27 | 15.12 ± 0.43 | 0.367 |
|  | INR | 1.03 ± 0.01 | 1.06 ± 0.02 | 0.303 |
|  | FIB (g/L) | 3.81 ± 0.21 | 4.29 ± 0.40 | 0.271 |
|  | D-dimer (mg/L) | 4.01 ± 0.60 | 3.07 ± 0.51 | 0.246 |

Comparisons of CCBs use *vs.* other antihypertensive medication using Student’s *t* test for normally distributed data and Fisher’s test was used for contingency data.

^a^ Data are presented as the mean ± SEM and n (%) for percentages. *P* <0.05 means statistically significant.

BMI, body mass index; SBP, systolic blood pressure; RBC, red blood cell; WBC, white blood cell; PLT, platelets; HB, hemoglobin; U-RBC, urine-red blood cell; U-WBC, urine-white blood cell, U-PRO, urine-protein; U-PH, urine-PH; ALT/GPT, glutamic pyruvic transaminase; AST/GOT, glutamic oxaloacetic transaminase; TBIL, total bilirubin; DBIL, direct bilirubin; IBIL, indirect bilirubin; APTT, activated partial thromboplastin time; PT, prothrombin time; TT, thrombin time; INR, international normalized ratio; FIB, fibrinogen.

**Extended Data Table 18. Measurements of aortic dissection detected by computed tomography imaging pre-TEVAR and operation records.**

| Number of patients | | CCBs use  (n=69) | Other antihypertensive medication  (n=26) | *P* value |
| --- | --- | --- | --- | --- |
| Aortic Dissection phase, n (%) | |  |  |  |
|  | Acute phase (≤ 14 days) | 46 (66.7%) | 18 (69.2%) | 0.812 |
|  | Subacute phase (14-90 days) | 21 (30.4%) | 8 (30.8%) | 1.000 |
|  | Chronic phase (≥ 90 days) | 2 (2.9%) | 0 | 1.000 |
| False lumen zone length grades ^a^ | | 6.00 ± 0.21 | 5.50 ± 0.33 | 0.203 |
| False lumen thrombus grades ^a, b^ | | 16.33 ± 0.68 | 15.08 ± 1.26 | 0.348 |
| Total tear numbers ^a^ | | 1.88 ±0.15 | 1.81 ±0.23 | 0.788 |
|  | Distal tear numbers | 0.82 ± 0.12 | 0.69 ± 0.17 | 0.559 |
| First intimal tear diameter (mm) ^a^ | | 9.00 ± 0.56 | 7.78 ± 0.93 | 0.296 |
| Diameter of aorta (mm) ^a^ | |  |  |  |
|  | Bottom of Zone 3 FL | 17.44 ± 0.94 | 15.83 ± 1.32 | 0.354 |
|  | Bottom of Zone 4 FL | 17.11 ± 0.88 | 17.79 ± 1.31 | 0.683 |
|  | DTA max FL | 21.14 ± 1.02 | 22.02 ± 1.84 | 0.663 |
| Volume of aorta (ml) ^a^ | |  |  |  |
|  | Zones 3-4 | 129.55 ± 8.29 | 127.45 ± 7.33 | 0.850 |
|  | Zone 5 | 92.40 ± 3.88 | 93.15 ± 4.90 | 0.915 |
|  | Zones 3-5 | 221.95 ± 11.58 | 220.60 ± 11.80 | 0.935 |
| Covered endograft length | | 179.62 ± 1.25 | 179.73 ±1.04 | 0.948 |

Comparisons of CCBs use *vs.* other antihypertensive medication using Student’s *t* test for normally distributed data and Fisher’s test was used for contingency data.

^a^ Data are presented as the mean ± SEM and n (%) for percentages. *P* <0.05 means statistically significant.

^b^ False lumen thrombus grades: 1, Complete thrombus; 2, Partial thrombus; 3, Patency of false lumen; Calculate zones 3-9 grades, respectively and plus all as the total false lumen thrombus grades.

TEVAR, thoracic endovascular aortic repair; FL, false lumen; DTA max, the max diameter of decreasing thoracic aorta.

**Extended Data Table 19. Characteristics of mice (AngII model) with CCBs and Sh_Scramble_, Sh*_Prkg1_* for 21 days.**

| Groups (♂) | AAV9-Sh_Scramble_ | | AAV9-Sh*_Prkg1_* | |
| --- | --- | --- | --- | --- |
|  | AngII + Vehicle | AngII + Verapamil | AngII + Vehicle | AngII + Verapamil |
|  | N=6 | N=16 | N=5 | N=16 |
| Weight -7 days (g) | 30.64 ± 0.30 | 31.36 ± 0.23 | 30.42 ± 0.42 | 30.87 ± 0.22 |
| Weight 0 day (g) | 30.89 ± 0.54 | 30.81 ± 0.42 | 30.69 ± 0.56 | 30.88 ± 0.36 |
| Weight 7 days (g) | 31.20 ± 0.57 | 30.00 ± 0.33 | 30.00 ± 0.97 | 31.23 ± 0.28 |
| Weight 14 days (g) | 30.58 ± 0.85 | 30.73 ± 0.40 | 30.54 ± 0.87 | 30.90 ± 0.32 |
| Weight 21 days (g) | 31.28 ± 0.48 | 31.93 ± 0.56 | 31.13 ± 0.63 | 31.40 ± 0.27 |
| SBP -14 days (mmHg) | 105.15 ± 0.96 | 104.24 ± 0.60 | 102.85 ± 1.56 | 104.73 ± 0.53 |
| SBP -7 days (mmHg) | 102.74 ± 1.75 | 104.83 ± 1.18 | 106.35 ± 3.52 | 105.09 ± 1.17 |
| SBP 0 days (mmHg) | 140.62 ± 0.79 | 141.48 ± 1.13 | 144.23 ± 3.52 | 143.00 ± 1.02 |
| SBP 7 days (mmHg) | 144.65 ± 1.76 | 113.54 ± 2.18* | 148.63 ± 2.61 | 118.47 ± 2.51* |
| SBP 14 days (mmHg) | 144.41 ± 1.41 | 112.52 ± 2.26* | 152.76 ± 3.67 | 122.06 ± 2.41* |
| SBP 21 days (mmHg) | 145.93 ± 3.35 | 114.45 ± 1.47* | 155.44 ± 3.14 | 127.42 ± 2.43* |
| TC (mg/dl) | 209.95 ± 19.94 | 189.57 ± 11.34^ns^ | 196.13 ± 20.49 | 204.26 ± 10.98^ns^ |
| TG (mg/dl) | 104.7 ± 10.27 | 99.57 ± 7.29^ns^ | 100.48 ± 5.72 | 94.91 ± 7.79^ns^ |

Each group weight was compared using Kruskal-Wallis test with Dunn's multiple comparisons test.

Comparison of AAV9-Sh_Scramble_ AngII + Vehicle group *vs.* AAV9-Sh_Scramble_ AngII + Verapamil group.

Comparison of AAV9-Sh*_Prkg1_* AngII + Vehicle group *vs.* AAV9-Sh*_Prkg1_* AngII + Verapamil group.

For SBP, Two-way ANOVA with Tukey multiple comparisons test.

For TC and TG, Kruskal-Wallis test with Dunn's multiple comparisons test.

Data were presented as mean ± SEM. **P* < 0.05; ns, not significant.

TC, total cholesterol; TG, triglyceride; SBP, systolic blood pressure.

**Extended Data Table 20. Characteristics of mice (BAPN model) with CCBs and Sh_Scramble_, Sh*_Prkg1_* for 21 days.**

| Groups (♂) | AAV9-Sh_Scramble_ | | AAV9-Sh*_Prkg1_* | |
| --- | --- | --- | --- | --- |
|  | BAPN + Vehicle | BAPN + Verapamil | BAPN + Vehicle | BPAN + Verapamil |
|  | N=20 | N=16 | N=10 | N=10 |
| Weight -7 days (g) | 14.15 ± 0.22 | 14.04 ± 0.26 | 14.97 ± 0.21 | 14.54 ± 0.27 |
| Weight 0 day (g) | 17.43 ± 0.35 | 17.37 ± 0.41 | 19.33 ± 0.22 | 18.82 ± 0.29 |
| Weight 7 days (g) | 18.93 ± 0.34 | 18.57 ± 0.44 | 21.19 ± 0.09 | 20.23 ± 0.37 |
| Weight 14 days (g) | 20.40 ± 0.39 | 19.72 ± 0.53 | 22.51 ± 0.27 | 21.21 ± 0.36 |
| Weight 21 days (g) | 21.00 ± 0.43 | 20.71 ± 0.74 | 23.18 ± 0.23 | 21.65 ± 0.42 |
| Water -7 days (ml/g/d) | 0.212 | 0.223 | 0.217 | 0.217 |
| Water 0 day (ml/g/d) | 0.209 | 0.219 | 0.201 | 0.191 |
| Water 7 days (ml/g/d) | 0.210 | 0.200 | 0.209 | 0.208 |
| Water 14 days (ml/g/d) | 0.222 | 0.218 | 0.216 | 0.212 |
| Water 21 days (ml/g/d) | 0.221 | 0.201 | 0.228 | 0.222 |

Each group weight was compared using Kruskal-Wallis test with Dunn's multiple comparisons test.

Weight data are presented as the mean ± SEM.

Water intake data were presented for per unit of body weight and per day.
